# A Multicenter Randomized Comparison of High-Intensity Interval Training and Moderate-Intensity Exercise to Recover Walking Post-Stroke: Results of the HIT-Stroke Trial

**DOI:** 10.1101/2022.08.01.22278246

**Authors:** Pierce Boyne, Sandra A. Billinger, Darcy S. Reisman, Oluwole O. Awosika, Sofia Buckley, Jamiah Burson, Daniel Carl, Matthew DeLange, Sarah Doren, Melinda Earnest, Myron Gerson, Madison Henry, Alli Horning, Jane Khoury, Brett Kissela, Abigail Laughlin, Kiersten McCartney, Thomas McQuaid, Allison Miller, Alexandra Moores, Jacqueline A. Palmer, Heidi Sucharew, Elizabeth Thompson, Erin Wagner, Jaimie Ward, Emily Wasik, Alicen A. Whitaker, Henry Wright, Kari Dunning

**Affiliations:** Department of Rehabilitation, Exercise and Nutrition Sciences, College of Allied Health Sciences, University of Cincinnati; Cincinnati, OH; Department of Neurology, School of Medicine, University of Kansas Medical Center; Kansas City, KS; Department of Cell Biology and Integrative Physiology, School of Medicine, University of Kansas Medical Center; Kansas City, KS; University of Kansas Alzheimer’s Research Disease Center, Fairway, KS; Department of Physical Medicine and Rehabilitation, School of Medicine, University of Kansas Medical Center; Kansas City, KS; Department of Physical Therapy, College of Health Sciences, University of Delaware; Newark, DE; Department of Neurology and Rehabilitation Medicine, College of Medicine, University of Cincinnati; Cincinnati, OH; Departments of Internal Medicine and Cardiology, College of Medicine, University of Cincinnati; Cincinnati, OH; Department of Physical Therapy, Rehabilitation Sciences, and Athletic Training, School of Health Professions, University of Kansas Medical Center; Kansas City, KS; Division of Biostatistics and Epidemiology, Cincinnati Children’s Hospital Medical Center; Cincinnati, OH; Department of Pediatrics, College of Medicine, University of Cincinnati; Cincinnati, OH

**Author notes:** **Corresponding author:** Pierce Boyne, PT, DPT, PhD, NCS, Health Sciences Building, room 267, 3225 Eden Ave., Cincinnati, OH, 45267-0394.

**Keywords:** locomotion, gait, aerobic exercise, HIIT

## Abstract

**Introduction:** For walking rehabilitation after stroke, training intensity and duration are critical dosing parameters that lack optimization. This trial aimed to determine the optimal training intensity (vigorous vs moderate) and minimum training duration (4, 8 or 12 weeks) needed to maximize immediate improvement in walking capacity in chronic stroke.

**Methods:** Persons with chronic post-stroke gait dysfunction at three centers were randomized to high-intensity interval training (HIT) or moderate intensity aerobic training (MAT), each involving 45 minutes of treadmill and overground walking exercise with a physical therapist, 3 times per week for 12 weeks. The HIT protocol used repeated 30 second bursts of walking at maximum safe speed, alternated with 30-60 second recovery periods, targeting an average aerobic intensity above 60% heart rate reserve (HRR). The MAT protocol used continuous walking with speed adjusted to maintain an initial target of 40 ± 5% HRR, progressing by 5% HRR every 2 weeks, up to 60% HRR as tolerated. Blinded assessment at baseline and after 4, 8 and 12 weeks of training included the 6-minute walk test (6MWT) as the primary measure of walking capacity.

**Results:** Randomized participants (N=55) attended 1,675 (85%) of 1,980 planned treatment sessions and 197 (90%) of 220 planned testing sessions. No serious adverse events related to study procedures occurred. Compared with MAT, HIT involved significantly higher training speeds (161% vs 96% baseline fastest 10-meter speed, p<0.0001) and mean aerobic intensity (61% vs 46% HRR, p<0.0001) across treatment visits. There was no significant between-group difference in 6MWT changes after 4 weeks of training (HIT +27 meters [95% CI: 6-48], MAT +12 meters [-9-33], p=0.28), but randomization to HIT resulted in significantly greater gains than MAT after 8 weeks (+58 [39-76] vs +29 [9-48] meters, p=0.02) and 12 weeks (+71 [49-94] vs +27 [3-50] meters, p=0.005) of training. HIT also showed significantly greater improvements than MAT on some measures of gait speed, fatigue and exercise capacity.

**Discussion:** These findings show proof of concept that vigorous training intensity is a critical dosing parameter for walking rehabilitation. In chronic stroke, vigorous walking exercise can produce significant and meaningful gains in walking capacity with only 4 weeks of training, but at least 12 weeks are needed to maximize immediate gains.

## INTRODUCTION

While the majority of stroke survivors eventually regain the ability to walk without physical assistance from another person, most in the chronic phase of recovery still lack sufficient speed and endurance to resume normal daily activities.^1-5^ The neuromotor impairment and aerobic deconditioning underlying these limitations in walking capacity can both be targeted simultaneously with locomotor exercise (i.e. task-specific walking practice at sufficient intensity to challenge the neuromotor and cardiopulmonary systems).^6-8^ With this intervention, training intensity appears to be a critical dosing parameter that strongly influences outcomes.^9-21^ However, the optimal intensity for improving walking recovery remains unknown.

Most randomized controlled studies of post-stroke locomotor exercise have tested moderate-intensity aerobic training (MAT), typically involving treadmill walking at average training heart rates (HR) between 40-60% heart rate reserve (HRR).^22-33^ These studies have found that MAT improves walking capacity and other outcomes significantly more than lower intensity walking practice or non-walking exercise. Consequently, this approach is currently recommended in stroke rehabilitation guidelines.^6-8^

Studies also suggest that a more vigorous training intensity (>60% HRR vs 40-60% HRR) could augment outcomes,^9^ but that a vigorous intensity can be difficult to achieve and sustain for many persons with stroke.^34-36^ Thus, high-intensity interval training (HIT) has emerged as a promising strategy for post-stroke locomotor exercise.^37,38^ This method involves bursts of fast walking alternated with recovery periods, and is designed to enable higher sustained intensities^39^ at lower perceived exertion than high-intensity continuous exercise.^40^ Initial stroke studies indicate that locomotor HIT is feasible, sufficiently safe for further study, can improve walking capacity and other outcomes,^15,16,41-47^ and has some early signs of potential for eliciting greater improvements than MAT.^43,44^

However, no prior studies have been designed to compare outcomes of HIT and MAT post-stroke. In addition, most prior HIT studies post-stroke used short 4-week training durations. Therefore, it was unknown whether a longer 12-week training duration typical of many stroke and cardiac rehabilitation trials^9,48^ could yield better outcomes.

The purpose of this multi-center trial was to determine the optimal training intensity (vigorous vs moderate) and the minimum training duration (4, 8 or 12 weeks) needed to maximize immediate improvement in walking capacity in chronic stroke. We hypothesized that: 1) 4 weeks of HIT would elicit significantly greater improvement in walking capacity compared to 4 weeks of MAT; and 2) Compared with 4 and 8 weeks of HIT, 12 weeks of HIT would elicit significantly greater improvements in walking capacity and increased benefit over MAT.

## METHODS

### Study overview

This study was approved by the University of Cincinnati IRB and performed in rehabilitation and exercise laboratories at the University of Cincinnati, University of Delaware and University of Kansas Medical Center with participant enrollment from January 2019 to April 2022. It was prospectively registered at ClinicalTrials.gov (NCT03760016) and a detailed protocol was published.^49^

### Multisite standardization

Procedures were standardized across sites using a detailed manual of procedures, video-based online personnel training, 2-day in-person training at each site and monthly web-meetings. This study also used direct-electronic data entry into REDCap forms that were programmed to provide real-time protocol reminders, automated calculations, prompts and feedback during each study visit. In addition, a software application used during each treatment visit was programmed to prompt training start/stop times and provide real-time intensity feedback.

### Participants

Participants were recruited from the community via outreach to clinicians and support groups, advertisement, existing databases and health record systems. Written informed consent was obtained prior to participation. Screening included a medical history and record review, physical examination, Fugl-Meyer lower limb motor function assessment,^50^ depression questionnaire,^51^ 10-meter walk test,^52,53^ treadmill acclimation and screening test,^52^ resting electrocardiogram (ECG) and a treadmill graded exercise test with ECG monitoring.^54^

#### Inclusion criteria were

1) Age 40-80 years at the time of consent; 2) Single stroke for which the participant sought treatment, 6 months to 5 years prior to consent; 3) Walking speed ≤ 1.0 m/s on the 10-meter walk test;^53,55^ 4) Able to walk 10m over ground with assistive devices as needed and no continuous physical assistance from another person;^56^ 5) Able to walk at least 3 minutes on the treadmill at ≥ 0.13 m/s (0.3 mph);^52^ 6) Stable cardiovascular condition (American Heart Association class B,^57^ allowing for exercise capacity <6 METs); and 7) Able to communicate with investigators, follow a 2-step command and correctly answer consent comprehension questions.

#### Exclusion criteria were

1) Exercise testing uninterpretable for ischemia or arrhythmia;^57^ 2) Evidence of significant arrhythmia or myocardial ischemia on treadmill ECG graded exercise test in the absence of recent (past year) more definitive clinical testing with negative result;^57^ 3) Hospitalization for cardiac or pulmonary disease within the past 3 months; 4) Implanted pacemaker or defibrillator; 5) Significant ataxia or neglect (score of 2 on NIH stroke scale item 7 or 11);^58^ 6) Severe lower limb spasticity (Ashworth >2);^59^ 7) Recent history (<3 months) of illicit drug or alcohol abuse or significant mental illness; 8) Major post-stroke depression (Patient Health Questionnaire [PHQ-9] ≥ 10)^51^ in the absence of depression management by a health care provider;^60^ 9) Currently participating in physical therapy or another interventional study; 10) Recent botulinum toxin injection to the paretic lower limb (<3 months) or planning to have lower limb botulinum toxin injection in the next 4 months; 11) Foot drop or lower limb joint instability without adequate stabilizing device, as assessed by a physical therapist; 12) Clinically significant neurologic disorder other than stroke; 13) Unable to walk outside the home prior to stroke; 14) Other significant medical condition likely to limit improvement or jeopardize safety as assessed by a physical therapist; 15) Pregnancy; and 16) Previous exposure to fast treadmill walking (>3 cumulative hours) during clinical or research therapy in the past year.

### Study design

Eligible participants were randomized to either HIT or MAT, with a target training volume of 45 minutes, 3 times per week for 12 weeks (Figure 1). This was divided into three 4-week (12-session) blocks separated by outcome testing. In each training block, up to one additional week was allowed to make up for missed sessions if needed. Outcomes were assessed by blinded raters before randomization (PRE) and after 4, 8 and 12 weeks of training (4WK, 8WK, 12WK), including measures of walking function, fatigue and aerobic capacity.

**Figure 1.**
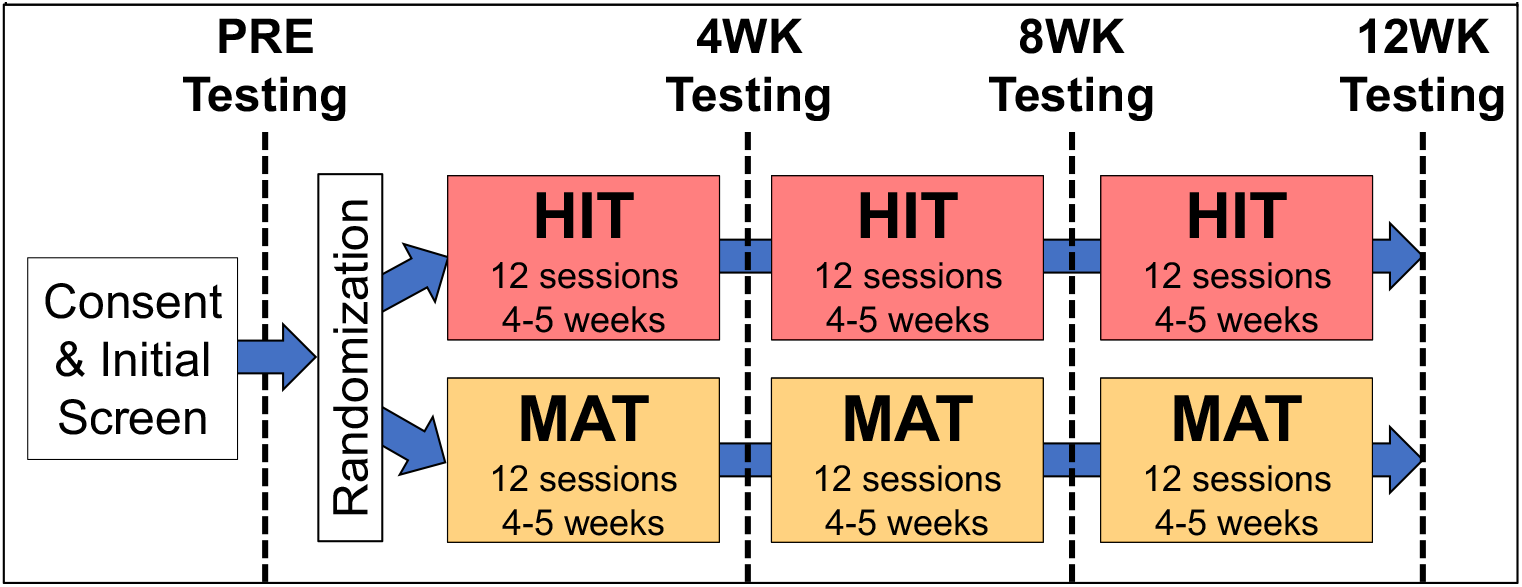
Study design. HIT, high-intensity interval training; MAT, moderate-intensity aerobic training; WK, week

### Randomization

After screening and PRE testing, eligible participants were randomized to HIT or MAT in a 1:1 ratio using the web-based REDCap randomization module.^61^ Randomization was stratified by site and baseline gait dysfunction (severe, <0.4 m/s; mild/moderate, 0.4-1.0 m/s),^62,63^ and used randomly-permuted block sizes of 2 or 4 to help ensure balanced groups while preventing personnel from being able to predict the last randomization within a block. The study statistician generated the random allocation sequence and kept it confidential from all other study personnel to ensure concealed allocation. When eligible participants arrived for the first training session, the treating therapist or study coordinator initiated the randomization within REDCap to irreversibly assign the participant to the next allocation in the randomized sequence before revealing the treatment assignment.

### Interventions

Training sessions were directed by a physical therapist with support from a research assistant as needed for data collection. Habitual orthotic devices were used and occasionally supplemented by additional orthotics to protect against ankle inversion sprain or severe knee hyperextension during training, based on the judgment of the treating therapist. Participants were not physically assisted with stepping but were guarded as needed and assisted for injury prevention during instances of severe gait instability or loss of balance. Participants who enrolled after the beginning of the COVID-19 pandemic wore personal protective equipment during testing and treatment sessions according to the infection control protocols at each site.

During overground training bouts, participants walked back and forth in a corridor. Habitual assistive devices were used during initial bouts, then therapists attempted to progress the participant to less restrictive device(s) if it better enabled achievement of the target intensity. When training on a treadmill (LiteGait Gaitkeeper 2200T, Mobility Research, Tempe, AZ, USA; or L8 Rehab Treadmill, Landice, Randolph, NJ, USA), participants wore a fall protection harness without any weight support (Balance Harness, MASS Rehab, Dayton, OH) connected to an overhead system (RehabMODULE, MASS Rehab, Dayton, OH, USA) and used a handrail connected to the fall protection system for balance support.^52^ This handrail was individually height-adjusted during treadmill training to allow upright walking posture.

Training intensity was monitored and recorded using Bluetooth Smart heart rate (HR) transmitters (H7 and H10, Polar Electro Oy, Kempele, Finland; or Rhythm24, Scosche Industries, Inc., Oxnard, CA, USA) wirelessly connected to an iPod touch (Apple, Cupertino, CA, USA) using the iCardio application (FitDigits, Inc., Ventura, CA, USA).^63,64^ The training protocols and individual prescribed target HR zones were programmed into the application to time each training bout, signal burst/recovery transitions (for HIT) and provide real-time intensity guidance. A forearm-worn iPod mount enabled hands-free monitoring for the treating therapist while guarding the participant.

Training HR zones were calculated using the HR reserve (HRR) method: (HR_peak_ – HR_rest_) x % HRR target + HR_rest_.^57^ During training, HR_peak_ was taken as the highest instantaneous HR obtained in any prior exercise testing session for that participant. HR_rest_ was measured at the beginning of each session in both sitting and standing. For this study, the HR_rest_ value in standing was used for target HR calculations. This was based on our pilot testing in which some participants with more severe deconditioning were found to have large HR increases just from standing that would have otherwise exceeded some of the target HR ranges and precluded walking practice if we had used the true HR_rest_ value for target HR calculation. Each session, the training protocol for both groups included: 1) a 3-minute warm-up of overground walking at a speed that increased HR to 30-40% HR reserve (HRR); 2) a 10-minute bout of overground HIT or MAT; 3) a 20-minute bout of treadmill HIT or MAT; 4) another 10-minute bout of overground HIT or MAT; and 5) a 2-minute cool down at 30-40% HRR (Figure 2).

**Figure 2.**
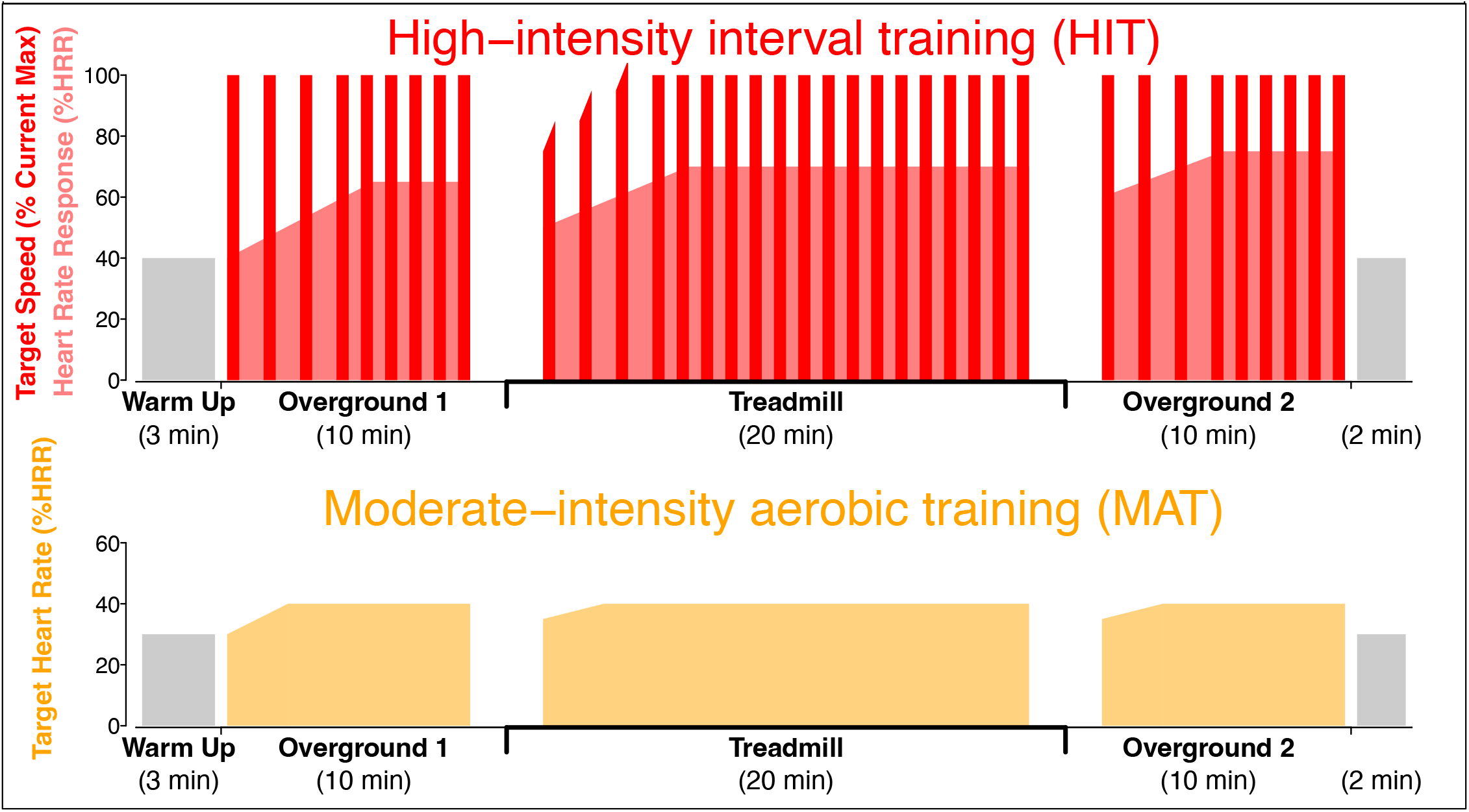
Training Protocol Schematics. HIT (upper panel) uses 30 second bursts at 100% maximum safe speed (dark red bars) alternated with 30-60 second passive recovery periods and within-session speed progression as able (not shown). This typically elicits a mean aerobic intensity >60% heart rate reserve (HRR; light red shading). The first three treadmill bursts depict example within-burst speed increases to find the current maximum safe treadmill speed. MAT (lower panel) uses continuous walking with speed adjusted as needed to maintain the moderate aerobic intensity target, which starts at 40% HRR (shown) and progresses by 5% HRR every 2 weeks, up to 60% HRR (not shown).

#### High-intensity interval training (HIT) protocol

The HIT group used a ‘short-interval’ HIT protocol that was specifically developed for locomotor exercise post-stroke.^43,63-65^ It involved repeated 30 second bursts of walking at maximum safe speed, alternated with 30-60 second passive recovery periods, targeting an average aerobic intensity above 60% HRR. During overground HIT, burst speed was maximized by placing cones or beanbags on the floor to mark the distance covered during each burst and encouraging participants to continually work towards making it past the marker before time ran out. During treadmill HIT, burst speed was selected to provide the maximum safe challenge for that participant and was progressed as able (or regressed as needed) throughout each session based on performance criteria to maintain constant challenge.^64^

#### Moderate-intensity aerobic training (MAT) protocol

Following current best-practice guidelines for stroke rehabilitation,^6,7^ the MAT group performed continuous walking practice with speed adjusted to maintain an initial target HR of 40 ± 5% HRR, progressing by 5% HRR every 2 weeks, up to 60% HRR as tolerated.^52^

### Treatment fidelity data collection

- *Training volume* was measured by the number of sessions attended, the number of treadmill and overground training bouts started and the total number of training minutes performed.
- *Neuromotor intensity* was measured by peak training speed in each bout. Overground speeds were captured using a stopwatch at the beginning and end of each overground training bout as the participants crossed over markings for the 10-meter walk test in the overground training corridor. Treadmill speeds were recorded from the treadmill display.
- *Aerobic intensity* was measured by HR data collected continuously by the Bluetooth HR monitors and iPod touch iCardio application during each training bout. These data were exported at 1/3 Hz and processed to calculate mean steady-state HR (excluding the first 3 minutes) and max HR for each bout as %HRR.
  ○ For real-time treatment monitoring, %HRR values were calculated using standing HR_rest_ and the most current HR_peak_, as described above. However, the use of standing HR_rest_ makes such %HRR values incomparable to previous studies and allowing HR_peak_ updates at 4WK or 8WK could distort %HRR comparisons across sessions and/or treatment groups. Therefore, to facilitate these comparisons, %HRR values were also calculated using true HR_rest_ (the smaller of the two HR_rest_ values obtained from each session, which was typically the sitting HR_rest_) and HR_peak_ from PRE testing only.
  ○ HR data preprocessing included spike-filtering and plotting to visually check for invalid data (signal dropout, residual non-physiologic spikes, episodes of supraventricular tachycardia and large elevations clearly attributed to coughing or laughing), while referencing notes and other data from each bout. Where possible, invalid data segments were filled using linear interpolation. When a bout began with invalid data, the first value at the beginning of the adjacent bout(s) in the same session was used to initialize the interpolation, to approximate the typical HR increase from a low starting value at the beginning of the bout. When a bout ended with invalid data, the final valid data point was carried forward, to approximate the typical steady mean trajectory at the end of the bout. However, when this did not reasonably approximate the likely mean HR trajectory, the HR data from that bout were discarded as invalid (e.g. when >50% of the HR data from a bout were invalid/missing or the mean trajectory was steeply sloped). If resting HR was missing or invalid for a session (e.g. due to an episode of supraventricular tachycardia that subsided during the session), it was imputed for HRR calculation by taking the mean resting HR across sessions for that participant.
- *Anaerobic intensity* was measured by blood lactate concentration after the treadmill training portion of the middle training session in each training week, using a finger stick and a point-of-care blood lactate analyzer (Lactate Plus, Nova Biomedical, Waltham, MA, USA).
- *Practice repetition* was measured by step count, recorded each session using a Stepwatch Activity Monitor (Modus Health, LLC, Edmonds, WA, USA) on the non-paretic ankle.^66^
- *Rating of perceived exertion* (RPE) for the session was measured by the Borg 6-20 scale^57^ at the end of each session.

### Adverse event (AE) monitoring

Each study visit, participants were asked about AEs in general then specifically queried about falls, injuries, pain and fatigue. During exercise testing and training, safety monitoring included HR, blood pressure, and continuous observation for other signs or symptoms of cardiorespiratory insufficiency, worsening neurologic impairments or orthopedic injury, using accepted stopping criteria.^57,67^ AEs were systematically categorized and graded using Common Terminology Criteria for Adverse Events,^68,69^ by a centralized physician blinded to group randomization, based on standardized information provided by the research team member who discovered the AE.

### Outcome assessment

Outcomes were assessed by blinded raters at PRE, 4WK, 8WK and 12WK. All measures were assessed in a single visit and were arranged in standardized order to mitigate fatigue effects whenever possible, with overground walking tests followed by seated questionnaires then treadmill exercise testing.^49^ Testing visits were scheduled 2-7 days after the last treatment visit in the preceding block and subsequent treatment blocks were scheduled to begin within 10 days of the last treatment visit. An individual participant was tested by the same rater at all time points whenever possible. Similar to training, participants used habitual assistive and orthotic devices during testing, wore a fall protection harness during treadmill walking and were not physically assisted but were guarded by a physical therapist and provided injury prevention assistance if needed.

#### Primary outcome measure

The primary outcome measure was walking capacity, measured by distance walked during the 6-minute walk test (6MWT).^33,70^ The 6MWT was the primary outcome measure because it can be influenced by both gait speed and aerobic fitness,^6,7^ the two primary factors contributing to limited walking capacity post-stroke and the two primary targets of HIT and MAT.^9^ Further, this test explains more variance in home and community ambulation than any other laboratory measure for persons with stroke,^71-73^ is a primary characteristic distinguishing between home and community ambulators,^74^ and is significantly associated with post-stroke quality of life.^75^ The 6MWT has also been accepted by the FDA as a primary outcome for registered trials across various neurologic and cardiopulmonary diagnoses.^76-80^ After stroke, it has excellent test-retest reliability (ICC: 0.97-0.99)^81-84^ and adequate inter-rater reliability (ICC: 0.78).^85^ The minimal clinically important difference is 20 meters and 50 meters is a large improvement.^86^ It was standardized according to guidelines,^70^ allowing minor common modifications to the testing course at each site.^49^

#### Secondary outcome measures

- Gait speed at self-selected and fastest speeds, measured by the 10-meter walk test.^52,53^
- Self-reported fatigue over the past 7 days, measured by the Patient Reported Outcomes Measurement Information System (PROMIS)-Fatigue Scale version 8a.^87,88^
- Aerobic capacity, measured by oxygen consumption rate (VO_2_) at the ventilatory threshold during a treadmill graded exercise test (GXT).^89^ The ventilatory threshold is a submaximal transition point that represents the upper intensity limit of prolonged aerobic activity,^90-92^ beyond which anaerobic metabolism becomes more dominant, limiting sustainability.^90,91,93^ Compared with peak VO_2_ during an exercise test, which is often confounded by motor impairment after stroke,^89,90,93,94^ the ventilatory threshold appears to provide a more specific measure of aerobic capacity in this population.^89^

#### Treadmill exercise testing

The treadmill GXT was done with ECG monitoring, blood pressure testing and gas exchange analysis, using a metabolic cart (TrueOne 2400, ParvoMedics, Salt Lake City, UT, USA) with a facemask interface. A blinded physical therapist monitored gait safety while additional study team member(s) monitored ECG, blood pressure and metabolic data. The symptom-limited GXT protocol was standardized to be 0.3 mph at 0% grade for the first 3 minutes, then to increase by 0.1 mph every 30 seconds. If a participant reached 3.5 mph, that speed was maintained, and the incline started increasing by 0.5% every 30 seconds. The test continued until the participant requested to stop, drifted backward on the treadmill and was unable to recover, had severe gait instability judged to pose an imminent safety risk, or reached a cardiovascular safety limit.^57^ At least 10 minutes after the GXT, participants also attempted a 3-minute verification test without respiratory gas collection to help ensure that the highest possible HR was reached to guide individualized training intensity prescription. This test was done at the peak successful speed and grade from the GXT or at a higher speed/grade if the blinded testing therapist judged that it would be feasible. It was stopped before 3 minutes if one of the GXT stop criteria occurred sooner.

#### Ventilatory threshold determination

Ventilatory threshold identification from the GXT metabolic data was done using a semi-automated method. First, breath-by-breath data from each test were resampled to 5 second averaging and truncated to the GXT duration, including VO_2_, the rate of carbon dioxide production (VCO_2_) and the expiratory air flow volume (VE). Excess CO_2_ was calculated as (VCO_2_^2^ / VO^2^) – VCO^2^, with all values in L/min, and the ventilatory equivalents of O_2_ and CO_2_ were calculated as VE/VO_2_ and VE/VCO_2_.^95-97^ Automated threshold detection was then applied separately to the excess CO_2_ and VE/VO_2_ time series^95-97^ using the R^98^ package ‘mcp’^99^ to identify the posterior distribution likelihood of the ventilatory threshold at each 5 second time point. This was performed separately after smoothing the data using 0, 10 and 20 second rolling averages. The median of the posterior distributions across all methods was taken as an automated point estimate for time to ventilatory threshold.

Two raters then independently verified or adjusted this automated estimate while blinded to group assignment. Raters viewed graphs of VO_2_, VCO_2_ and VE by time, excess CO_2_ by time, VE/VO_2_ and VE/VCO_2_ by time and VCO_2_ by VO_2_ (V slope), across all testing timepoints for each participant. Automated posterior distribution likelihoods and point estimates were also displayed on the graphs. The ventilatory threshold was identified by an upward break point in the excess CO_2_ and VE/VO_2_ data, disproportionate to any VE/VCO_2_ increase, where the slope of the VCO_2_ by VO_2_ relationship increased above 1.^95-97^ If no threshold occurred during the test, time to threshold was set at the end of the GXT. Inter-rater reliability was excellent (ICC2,1: 0.98 [95%CI: 0.97-0.99], mean absolute error: 0.47 ± 0.87 minutes) despite a small systematic difference in mean time-to-threshold between raters (10.2 ± 4.9 versus 10.5 ± 4.7 minutes, p<0.0001). Final time-to-threshold values were determined by agreement (10.4 ± 4.7 minutes) and VO_2_ at the ventilatory threshold was calculated by averaging the VO_2_ data in a 20 second window around this time point.

#### Other exercise test measures to facilitate interpretation and exploratory outcomes

- Exercise capacity (not necessarily specific to aerobic fitness) was measured by GXT duration, peak GXT speed, peak GXT VO_2_, peak GXT HR and HR_peak_ (highest HR between GXT and verification test). Peak VO_2_ was calculated from 5 second averaged data after smoothing with a 20 second rolling average. Peak HR was the instantaneous peak from the ECG data.
- To facilitate interpretation of whether maximal *aerobic* capacity was reached during exercise testing:
  - HR_peak_ was also expressed as % age-predicted maximal HR, calculated using 164 – (0.7 x age [years])^100^ for participants taking a β-blocker medication and 206.9 – (0.67 x age [years])^101^ otherwise. Values are typically ≥90% at maximal aerobic capacity.^57^
  - Peak respiratory exchange ratio (VCO_2_/VO_2_) was calculated, using the same preprocessing as peak VO_2_. Values are typically at least ≥1.05 at maximal aerobic capacity.^57^
  - Rating of perceived exertion (Borg 6-20) was obtained at the end of the GXT. Values are typically ≥17 at maximal aerobic capacity.^57^
- Treadmill speed at the ventilatory threshold was measured to facilitate interpretation of any changes in VO_2_ at the ventilatory threshold. This represents the fastest speed that should have prolonged sustainability without accumulating anaerobic metabolites. It was calculated by averaging the protocol speed values in a 20 second window around the ventilatory threshold time.
- Metabolic cost of treadmill gait [mLO_2_/kg/m] during the GXT was calculated as VO_2_ [mL/kg/min] / speed [m/min], where lower values represent more efficient gait.^102^ Values were averaged in the last 3 minutes of each GXT. However, since faster speeds are typically more efficient after stroke,^102^ we also averaged the metabolic cost data in the last 3 minutes of the *shortest* test for each participant. This matched the speeds across time points to assess how much any changes in efficiency were related to (or independent from) any changes in speed.^103^
- Gait stability was measured with the functional ambulation category,^104^ based on participant performance during the 10-meter and 6-minute walk tests. Since we did not observe participants walking on nonlevel surfaces, the highest category (independent on nonlevel surfaces) was collapsed into the second highest category (independent on level surfaces), yielding a score range from 0-4+.
- Perceived balance confidence was measured with the activities-specific balance confidence (ABC) scale, a questionnaire with 16 items averaged to yield a total confidence score from 0-100%.^105,106^
- Quality of life related to mobility, self-care, usual activities, pain/discomfort and anxiety/depression were measured with the EuroQOL-5D-5L questionnaire.^107,108^ A total ‘misery score’ was calculated by summing the 5 item scores.^108^ Individual item scores were also analyzed for mobility and usual activities, given that these constructs were targeted by the interventions. Higher values represent worse problems for each of these scores.

### Statistical analysis

Analyses followed a prespecified plan,^49^ used an *intent-to-treat* approach and were done with SAS^®^, version 9.4 (SAS Institute, Cary, NC, USA). The study statistician remained blinded to treatment groups until after the primary analysis. To assess randomization performance, baseline participant characteristics were compared between groups with independent t-tests and Fisher exact tests. Baseline data were also expressed as a percentage of normative predicted values for self-selected gait speed,^55^ 6MWT distance,^109^ VO_2_ at the ventilatory threshold^92,110^ and VO_2_-peak.^110^ The HR_peak_ increase attributable to the verification test was quantified by comparing the GXT peak HR to HR_peak_ with a paired t-test.

#### Treatment fidelity analysis

Training intensity, repetition and perceived exertion data were compared between groups to evaluate treatment fidelity. Each of these dependent variables was tested in a separate statistical model, which included fixed effects for treatment group, session number (modeled as a categorical effect), training bout (for variables collected at overground 1, treadmill and overground 2 each session) and all possible interactions. Repeated sessions within the same participant were modeled with compound symmetry covariance and repeated bouts within the same session were modeled with unconstrained covariance. Variances were not constrained to be the same between sessions or between treatment groups. Missing data were handled with the method of maximum likelihood.

#### Adverse event (AE) analysis

To assess the relative odds of harms, post-randomization AEs were compared between treatment groups with logistic regression. Separate models were tested for overall AEs and for each AE categorization, using the number of participants with an AE in that category as the dependent variable and fixed effects for [treatment group], [study site] and [baseline gait dysfunction (<0.4, 0.4-1.0 m/s)]. If only one group had AE(s), a continuity correction was added of 0.5 AEs to each group to permit calculation of the odds ratio. In this case, it was not possible to adjust for study site or baseline dysfunction.

#### Hypothesis testing

Primary hypothesis testing used a linear model with the 6MWT as the dependent variable and fixed effects for treatment group, testing time point (PRE, 4WK, 8WK, POST), group by time interaction, study site, study site by time interaction, baseline gait dysfunction and baseline gait dysfunction by time interaction, with unconstrained covariance between repeated testing time points within the same participant and a significance threshold of p<0.05. Secondary outcomes were tested using the same model, with false discovery rate (FDR) correction^111^ to the significance threshold to control for multiple testing time points (4WKΔ, 8WKΔ, 12WKΔ) and the 5 primary or secondary outcome measures. Other exercise test measures and exploratory outcomes were tested with FDR correction across testing time points and all tested measures. These analyses handled any missing data with the method of maximum likelihood, which assumes that data were missing at random.^112^

#### Analysis stratified by baseline gait dysfunction subgroup

To preliminarily examine whether the primary results differed for participants with severe versus mild/moderate baseline gait dysfunction, we added a baseline gait dysfunction by group by time interaction effect to the primary model. In chronic stroke, persons with mild/moderate gait dysfunction have been found to have better responsiveness to locomotor exercise.^62,63^

#### Sensitivity analysis for missing data assumptions

While the ‘missing at random’ assumption is common in the analysis of clinical trials, it is not testable and could be violated in many typical circumstances, like when outcome data are missing because of AE-related participant withdrawal.^112,113^ To assess how much the results depended on the ‘missing at random’ assumption, we repeated the primary analysis assuming that participants with missing outcomes due to AE-related withdrawal had poor outcomes, similar to Duncan and colleagues.^60^ For any outcome data point missing because of AE-related withdrawal, the true value was assumed to be distributed around either the baseline or the last observation for that participant, whichever was smaller. If the data point missingness was not AE-related, the true value was assumed to be distributed around the last observation for that participant. These distributions for the true values were assumed to be normal with a standard deviation of 15 m, matching the observed standard deviation of 6MWT changes after 4 weeks of no intervention in a similar population.^63^ Using the general multiple imputation framework,^112,113^ 50 datasets were generated by random sampling to impute the missing values, each dataset was analyzed, and the model estimates were pooled across datasets.

### Sample size

This study was powered to detect a between-group difference of 20 m in 6MWT change^86^, using the software ‘GLIMMPSE’.^114^ We estimated that the MAT group would improve by 15 m every 4 weeks^43^ and set the HIT estimate 20 m larger at each time point. Variance & covariance parameters were estimated by pooling data across our prior 4-week pilot studies and extrapolating the repeated measures correlations involving the later 8WK and 12WK time points using the highest suggested exponential decay rate (0.5).^114^ These calculations indicated a total target sample size of 40 for ≥80% power at a two-sided significance threshold of 0.05. To account for up to 20% attrition, we initially planned to enroll and randomize 50 participants. However, after having to withdraw four participants directly due to COVID-19 shutdown, we opted to increase the enrollment target to 55. This decision was made before any analysis of outcome data.

## RESULTS

### Recruitment and retention

We consented and screened 74 participants to find 55 who were eligible for enrollment and randomized (Figure 3). No baseline characteristic significantly differed between groups (Table 1). In total, participants attended 1,675 (85%) of 1,980 planned treatment sessions and 197 (90%) of 220 planned testing sessions.

**Table 1.**
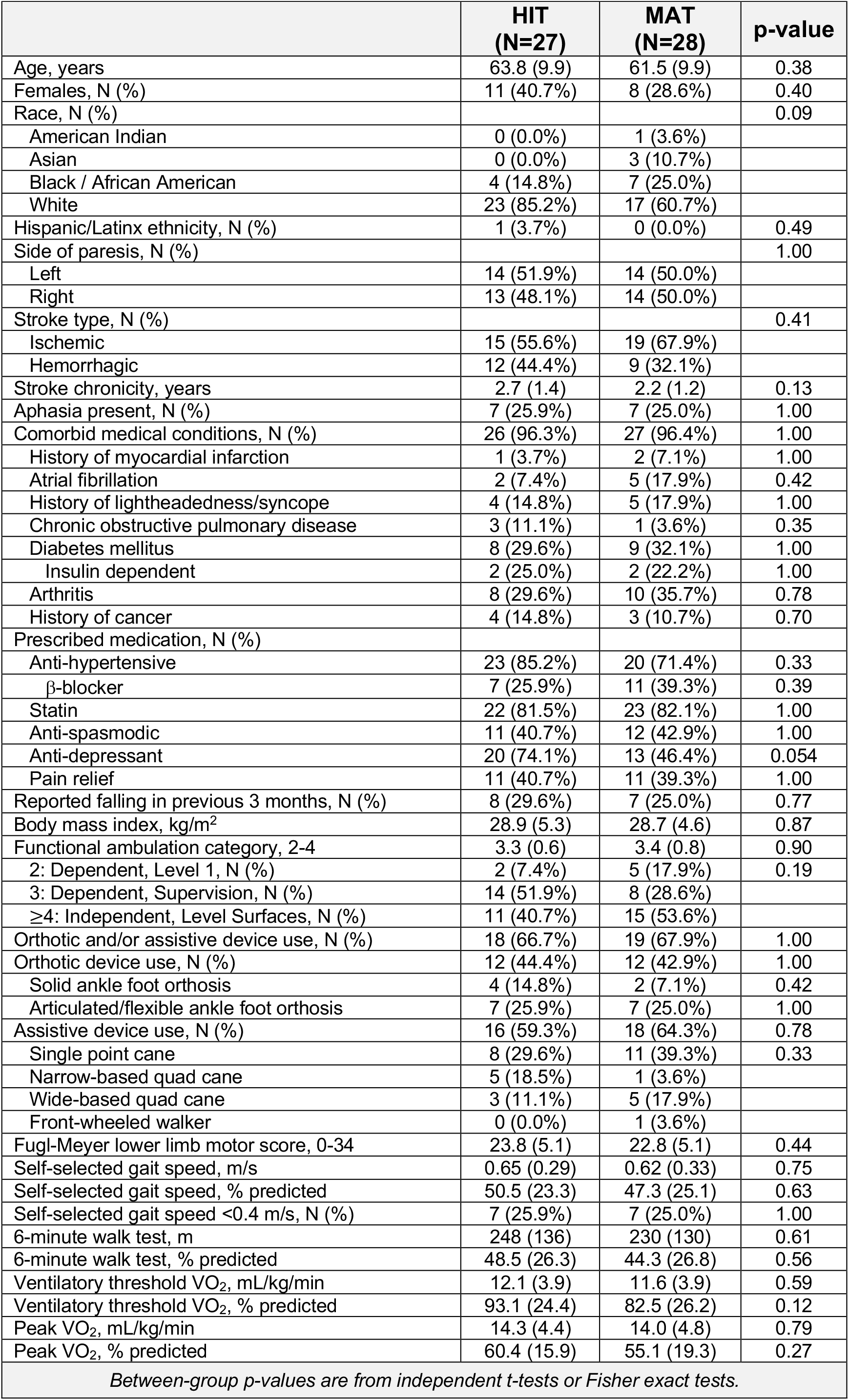
Baseline Participant Characteristics. *Values are mean (SD) or N (%)*.

**Figure 3.**
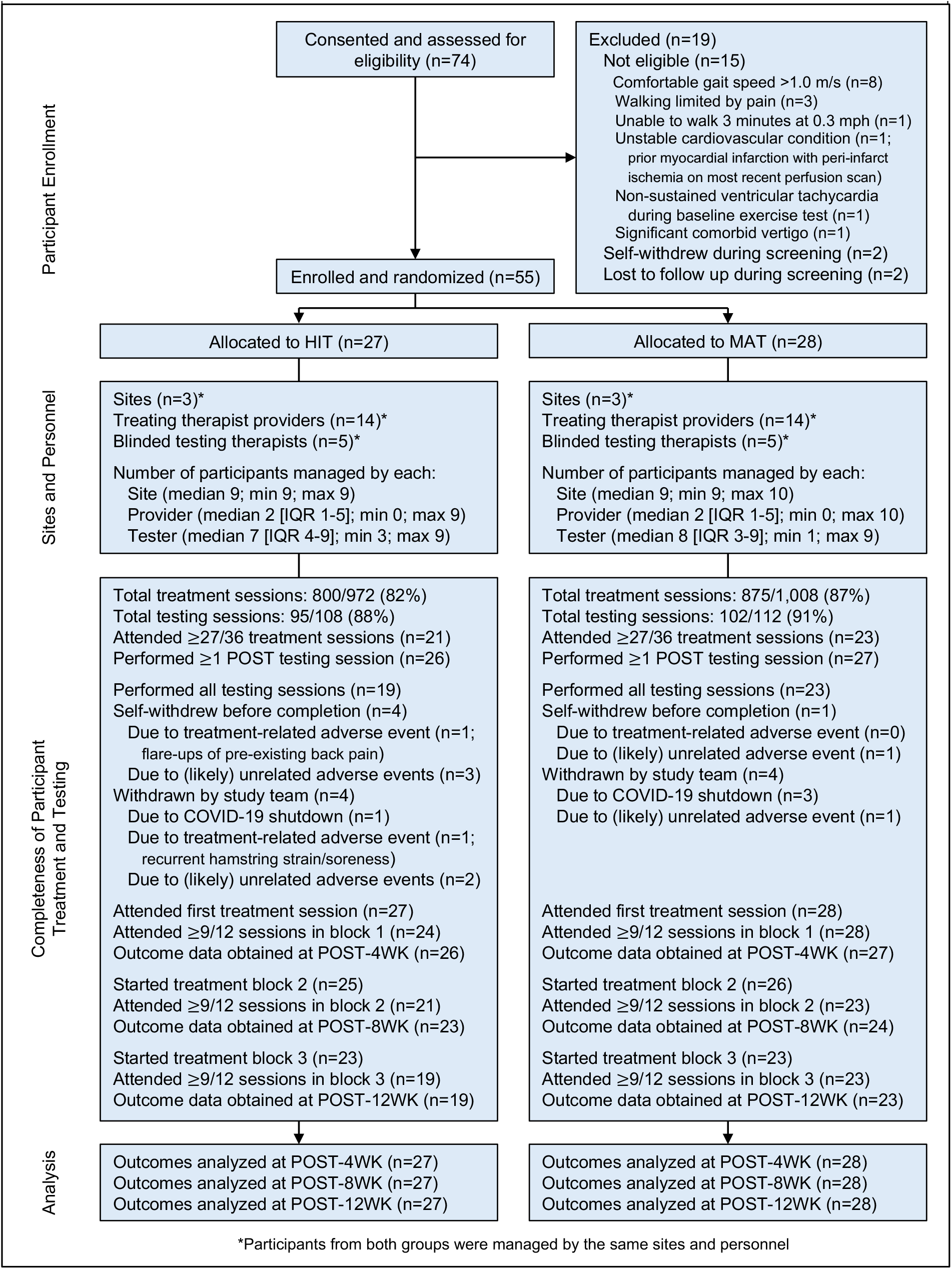
CONSORT flow diagram. *Participants from both groups were managed by the same sites and personnel

### Treatment fidelity

In total during the attended treatment sessions, participants initiated 5,006 (∼100%) of 5,025 planned overground and treadmill bouts (HIT, 2,383/2,400 [99%]; MAT, 2,623/2,625 [∼100%]), and performed 66,598 (99%) of 67,000 planned training minutes (HIT, 31,662/32,000 [99%]; MAT, 34,936/35,000 [∼100%]), including intermittent rest breaks.

- Peak training speed was recorded during 4,970 (99%) of the 5,006 initiated bouts (HIT, 2,364/2,383 [99%]; MAT, 2,606/2623 [99%]). Compared with MAT, HIT involved significantly faster training speed during all bouts and training blocks (Figure 4; Table 2). For HIT, training speed was significantly faster on the treadmill versus overground. Conversely for MAT, speed was significantly faster overground versus the treadmill. Training speed significantly increased across sessions for both protocols.

**Table 2.**
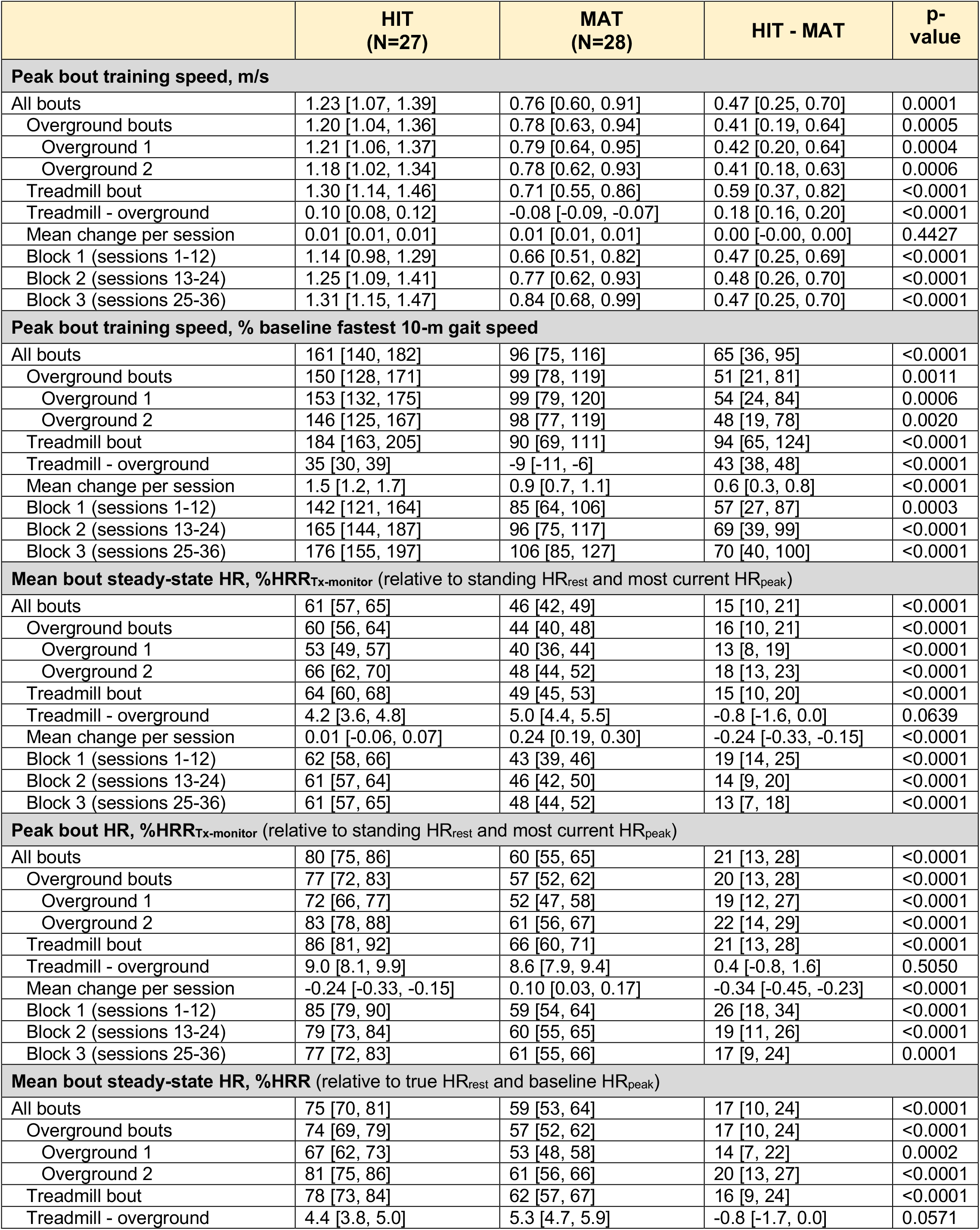
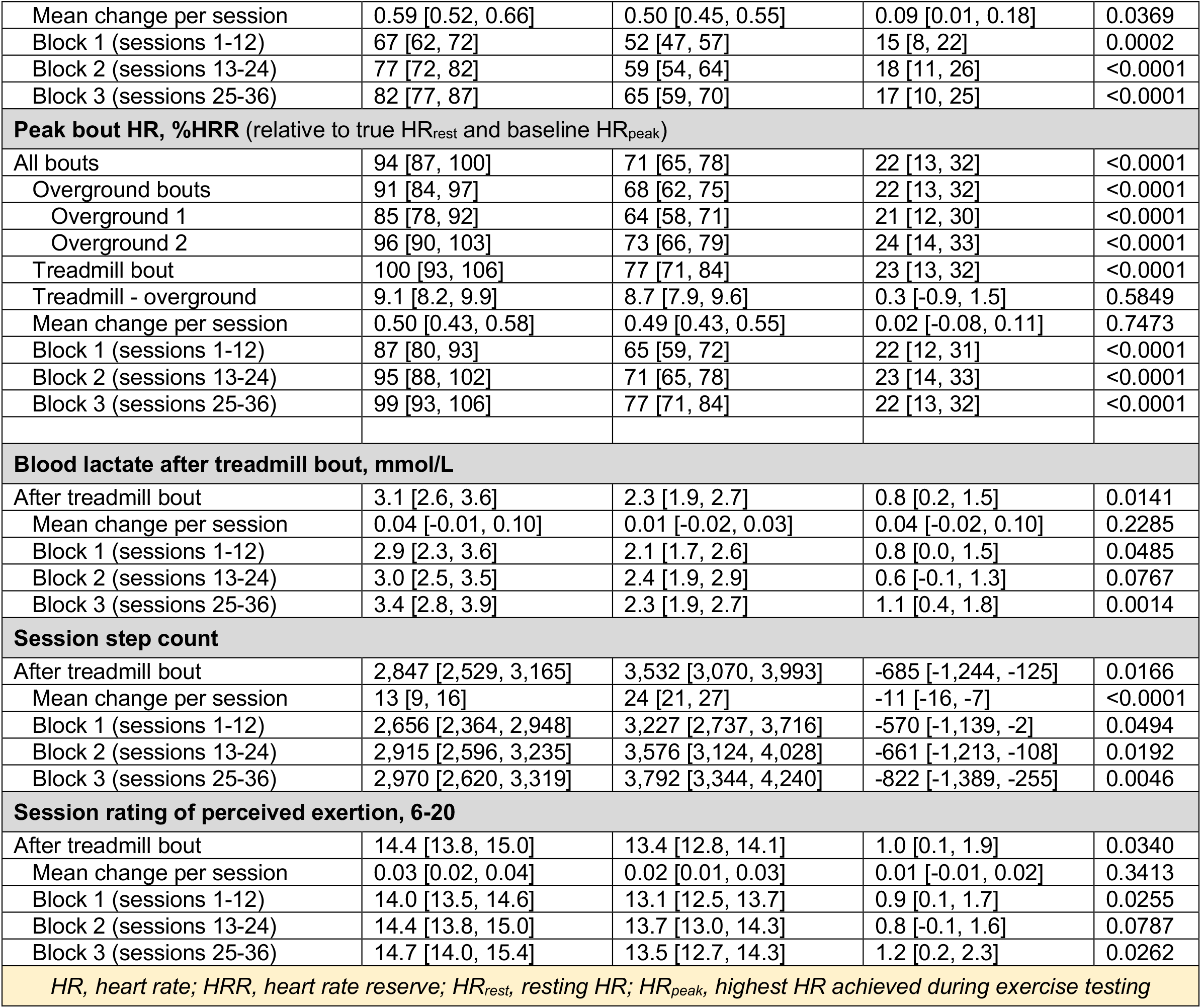
Treatment Intensity. Values are model estimates [95% CI]. Speed data are peak values within a bout. Heart rate data are presented as steady-state mean (excluding first 3 minutes) or peak values within a bout. Lactate, step counts and perceived exertion data are singular session values. All data are averaged across sessions and participants.

|  | HIT<br>(N=27) | MAT<br>(N=28) | HIT - MAT | p-value |
| --- | --- | --- | --- | --- |
| **Peak bout training speed, m/s** |  |  |  |  |
| All bouts | 1.23 [1.07, 1.39] | 0.76 [0.60, 0.91] | 0.47 [0.25, 0.70] | 0.0001 |
| Overground bouts | 1.20 [1.04, 1.36] | 0.78 [0.63, 0.94] | 0.41 [0.19, 0.64] | 0.0005 |
| Overground 1 | 1.21 [1.06, 1.37] | 0.79 [0.64, 0.95] | 0.42 [0.20, 0.64] | 0.0004 |
| Overground 2 | 1.18 [1.02, 1.34] | 0.78 [0.62, 0.93] | 0.41 [0.18, 0.63] | 0.0006 |
| Treadmill bout | 1.30 [1.14, 1.46] | 0.71 [0.55, 0.86] | 0.59 [0.37, 0.82] | <0.0001 |
| Treadmill - overground | 0.10 [0.08, 0.12] | -0.08 [-0.09, -0.07] | 0.18 [0.16, 0.20] | <0.0001 |
| Mean change per session | 0.01 [0.01, 0.01] | 0.01 [0.01, 0.01] | 0.00 [-0.00, 0.00] | 0.4427 |
| Block 1 (sessions 1-12) | 1.14 [0.98, 1.29] | 0.66 [0.51, 0.82] | 0.47 [0.25, 0.69] | <0.0001 |
| Block 2 (sessions 13-24) | 1.25 [1.09, 1.41] | 0.77 [0.62, 0.93] | 0.48 [0.26, 0.70] | <0.0001 |
| Block 3 (sessions 25-36) | 1.31 [1.15, 1.47] | 0.84 [0.68, 0.99] | 0.47 [0.25, 0.70] | <0.0001 |
| **Peak bout training speed, % baseline fastest 10-m gait speed** |  |  |  |  |
| All bouts | 161 [140, 182] | 96 [75, 116] | 65 [36, 95] | <0.0001 |
| Overground bouts | 150 [128, 171] | 99 [78, 119] | 51 [21, 81] | 0.0011 |
| Overground 1 | 153 [132, 175] | 99 [79, 120] | 54 [24, 84] | 0.0006 |
| Overground 2 | 146 [125, 167] | 98 [77, 119] | 48 [19, 78] | 0.0020 |
| Treadmill bout | 184 [163, 205] | 90 [69, 111] | 94 [65, 124] | <0.0001 |
| Treadmill - overground | 35 [30, 39] | -9 [-11, -6] | 43 [38, 48] | <0.0001 |
| Mean change per session | 1.5 [1.2, 1.7] | 0.9 [0.7, 1.1] | 0.6 [0.3, 0.8] | <0.0001 |
| Block 1 (sessions 1-12) | 142 [121, 164] | 85 [64, 106] | 57 [27, 87] | 0.0003 |
| Block 2 (sessions 13-24) | 165 [144, 187] | 96 [75, 117] | 69 [39, 99] | <0.0001 |
| Block 3 (sessions 25-36) | 176 [155, 197] | 106 [85, 127] | 70 [40, 100] | <0.0001 |
| **Mean bout steady-state HR, %HRR<sub>Tx-monitor</sub> (relative to standing HR<sub>rest</sub> and most current HR<sub>peak</sub>)** |  |  |  |  |
| All bouts | 61 [57, 65] | 46 [42, 49] | 15 [10, 21] | <0.0001 |
| Overground bouts | 60 [56, 64] | 44 [40, 48] | 16 [10, 21] | <0.0001 |
| Overground 1 | 53 [49, 57] | 40 [36, 44] | 13 [8, 19] | <0.0001 |
| Overground 2 | 66 [62, 70] | 48 [44, 52] | 18 [13, 23] | <0.0001 |
| Treadmill bout | 64 [60, 68] | 49 [45, 53] | 15 [10, 20] | <0.0001 |
| Treadmill - overground | 4.2 [3.6, 4.8] | 5.0 [4.4, 5.5] | -0.8 [-1.6, 0.0] | 0.0639 |
| Mean change per session | 0.01 [-0.06, 0.07] | 0.24 [0.19, 0.30] | -0.24 [-0.33, -0.15] | <0.0001 |
| Block 1 (sessions 1-12) | 62 [58, 66] | 43 [39, 46] | 19 [14, 25] | <0.0001 |
| Block 2 (sessions 13-24) | 61 [57, 64] | 46 [42, 50] | 14 [9, 20] | <0.0001 |
| Block 3 (sessions 25-36) | 61 [57, 65] | 48 [44, 52] | 13 [7, 18] | <0.0001 |
| **Peak bout HR, %HRR<sub>Tx-monitor</sub> (relative to standing HR<sub>rest</sub> and most current HR<sub>peak</sub>)** |  |  |  |  |
| All bouts | 80 [75, 86] | 60 [55, 65] | 21 [13, 28] | <0.0001 |
| Overground bouts | 77 [72, 83] | 57 [52, 62] | 20 [13, 28] | <0.0001 |
| Overground 1 | 72 [66, 77] | 52 [47, 58] | 19 [12, 27] | <0.0001 |
| Overground 2 | 83 [78, 88] | 61 [56, 67] | 22 [14, 29] | <0.0001 |
| Treadmill bout | 86 [81, 92] | 66 [60, 71] | 21 [13, 28] | <0.0001 |
| Treadmill - overground | 9.0 [8.1, 9.9] | 8.6 [7.9, 9.4] | 0.4 [-0.8, 1.6] | 0.5050 |
| Mean change per session | -0.24 [-0.33, -0.15] | 0.10 [0.03, 0.17] | -0.34 [-0.45, -0.23] | <0.0001 |
| Block 1 (sessions 1-12) | 85 [79, 90] | 59 [54, 64] | 26 [18, 34] | <0.0001 |
| Block 2 (sessions 13-24) | 79 [73, 84] | 60 [55, 65] | 19 [11, 26] | <0.0001 |
| Block 3 (sessions 25-36) | 77 [72, 83] | 61 [55, 66] | 17 [9, 24] | 0.0001 |
| **Mean bout steady-state HR, %HRR (relative to true HR<sub>rest</sub> and baseline HR<sub>peak</sub>)** |  |  |  |  |
| All bouts | 75 [70, 81] | 59 [53, 64] | 17 [10, 24] | <0.0001 |
| Overground bouts | 74 [69, 79] | 57 [52, 62] | 17 [10, 24] | <0.0001 |
| Overground 1 | 67 [62, 73] | 53 [48, 58] | 14 [7, 22] | 0.0002 |
| Overground 2 | 81 [75, 86] | 61 [56, 66] | 20 [13, 27] | <0.0001 |
| Treadmill bout | 78 [73, 84] | 62 [57, 67] | 16 [9, 24] | <0.0001 |
| Treadmill - overground | 4.4 [3.8, 5.0] | 5.3 [4.7, 5.9] | -0.8 [-1.7, 0.0] | 0.0571 |
| Mean change per session | 0.59 [0.52, 0.66] | 0.50 [0.45, 0.55] | 0.09 [0.01, 0.18] | 0.0369 |
| Block 1 (sessions 1-12) | 67 [62, 72] | 52 [47, 57] | 15 [8, 22] | 0.0002 |
| Block 2 (sessions 13-24) | 77 [72, 82] | 59 [54, 64] | 18 [11, 26] | <0.0001 |
| Block 3 (sessions 25-36) | 82 [77, 87] | 65 [59, 70] | 17 [10, 25] | <0.0001 |
| **Peak bout HR, %HRR (relative to true HR<sub>rest</sub> and baseline HR<sub>peak</sub>)** |  |  |  |  |
| All bouts | 94 [87, 100] | 71 [65, 78] | 22 [13, 32] | <0.0001 |
| Overground bouts | 91 [84, 97] | 68 [62, 75] | 22 [13, 32] | <0.0001 |
| Overground 1 | 85 [78, 92] | 64 [58, 71] | 21 [12, 30] | <0.0001 |
| Overground 2 | 96 [90, 103] | 73 [66, 79] | 24 [14, 33] | <0.0001 |
| Treadmill bout | 100 [93, 106] | 77 [71, 84] | 23 [13, 32] | <0.0001 |
| Treadmill - overground | 9.1 [8.2, 9.9] | 8.7 [7.9, 9.6] | 0.3 [-0.9, 1.5] | 0.5849 |
| Mean change per session | 0.50 [0.43, 0.58] | 0.49 [0.43, 0.55] | 0.02 [-0.08, 0.11] | 0.7473 |
| Block 1 (sessions 1-12) | 87 [80, 93] | 65 [59, 72] | 22 [12, 31] | <0.0001 |
| Block 2 (sessions 13-24) | 95 [88, 102] | 71 [65, 78] | 23 [14, 33] | <0.0001 |
| Block 3 (sessions 25-36) | 99 [93, 106] | 77 [71, 84] | 22 [13, 32] | <0.0001 |
| **Blood lactate after treadmill bout, mmol/L** |  |  |  |  |
| After treadmill bout | 3.1 [2.6, 3.6] | 2.3 [1.9, 2.7] | 0.8 [0.2, 1.5] | 0.0141 |
| Mean change per session | 0.04 [-0.01, 0.10] | 0.01 [-0.02, 0.03] | 0.04 [-0.02, 0.10] | 0.2285 |
| Block 1 (sessions 1-12) | 2.9 [2.3, 3.6] | 2.1 [1.7, 2.6] | 0.8 [0.0, 1.5] | 0.0485 |
| Block 2 (sessions 13-24) | 3.0 [2.5, 3.5] | 2.4 [1.9, 2.9] | 0.6 [-0.1, 1.3] | 0.0767 |
| Block 3 (sessions 25-36) | 3.4 [2.8, 3.9] | 2.3 [1.9, 2.7] | 1.1 [0.4, 1.8] | 0.0014 |
| **Session step count** |  |  |  |  |
| After treadmill bout | 2,847 [2,529, 3,165] | 3,532 [3,070, 3,993] | -685 [-1,244, -125] | 0.0166 |
| Mean change per session | 13 [9, 16] | 24 [21, 27] | -11 [-16, -7] | <0.0001 |
| Block 1 (sessions 1-12) | 2,656 [2,364, 2,948] | 3,227 [2,737, 3,716] | -570 [-1,139, -2] | 0.0494 |
| Block 2 (sessions 13-24) | 2,915 [2,596, 3,235] | 3,576 [3,124, 4,028] | -661 [-1,213, -108] | 0.0192 |
| Block 3 (sessions 25-36) | 2,970 [2,620, 3,319] | 3,792 [3,344, 4,240] | -822 [-1,389, -255] | 0.0046 |
| **Session rating of perceived exertion, 6-20** |  |  |  |  |
| After treadmill bout | 14.4 [13.8, 15.0] | 13.4 [12.8, 14.1] | 1.0 [0.1, 1.9] | 0.0340 |
| Mean change per session | 0.03 [0.02, 0.04] | 0.02 [0.01, 0.03] | 0.01 [-0.01, 0.02] | 0.3413 |
| Block 1 (sessions 1-12) | 14.0 [13.5, 14.6] | 13.1 [12.5, 13.7] | 0.9 [0.1, 1.7] | 0.0255 |
| Block 2 (sessions 13-24) | 14.4 [13.8, 15.0] | 13.7 [13.0, 14.3] | 0.8 [-0.1, 1.6] | 0.0787 |
| Block 3 (sessions 25-36) | 14.7 [14.0, 15.4] | 13.5 [12.7, 14.3] | 1.2 [0.2, 2.3] | 0.0262 |
| *HR, heart rate; HRR, heart rate reserve; HR<sub>rest</sub>, resting HR; HR<sub>peak</sub>, highest HR achieved during exercise testing* |  |  |  |  |

**Figure 4.**
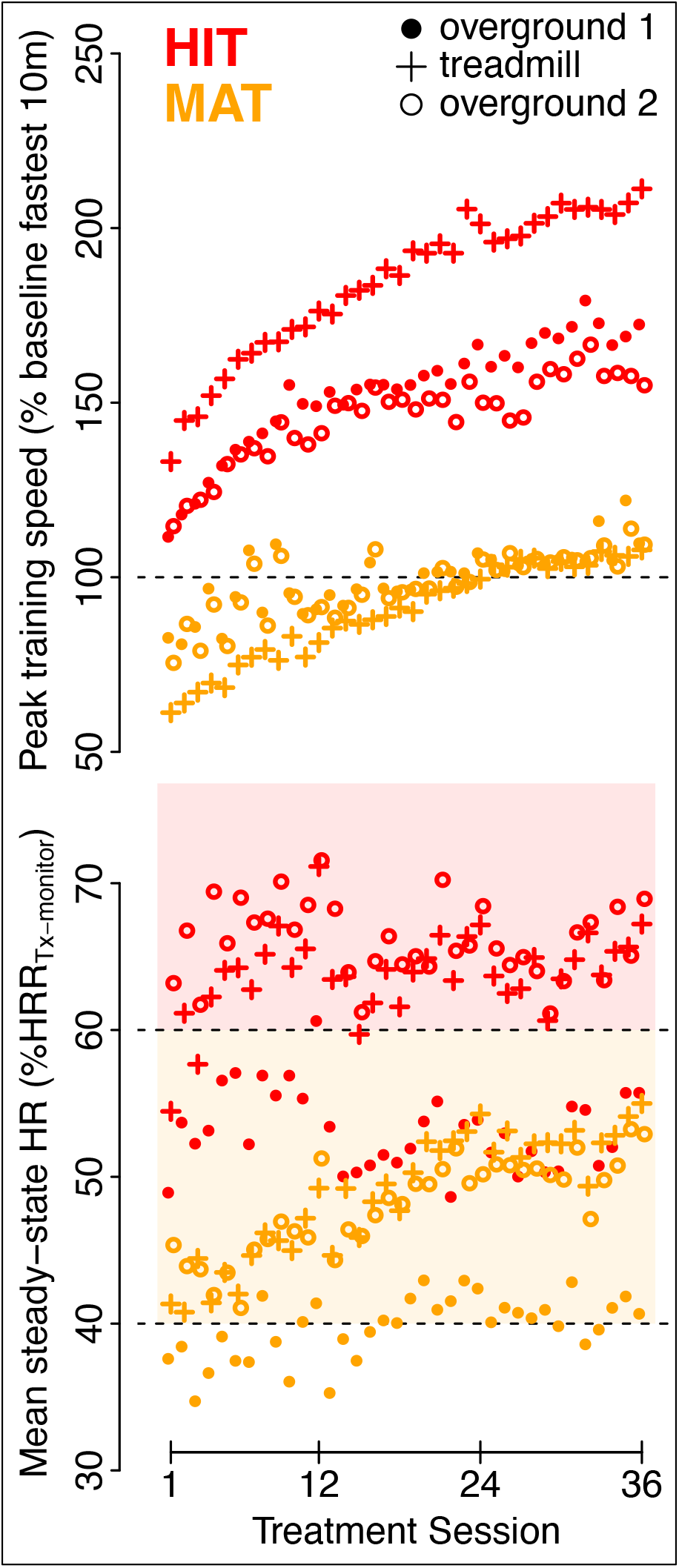
Treatment intensity. Values are model estimates for each bout of each session, averaging data across participants. Upper panel: Speed data are peak values within a bout. Lower panel: Steady-state mean heart rate (HR) excludes the first 3 minutes of the bout. For treatment (Tx) monitoring, % HR reserve (HRR) was relative to standing HR_rest_ and the most current HR_peak_, which could increase after sessions 12 and 24. Shading shows aerobic intensity zones: light orange, moderate; light red, vigorous
- Valid HR recordings were obtained for 65,666 (99%) of the 66,598 performed training minutes (HIT, 31,211/31,662 [99%]); MAT, 34,456/34,936 [99%]). Compared with MAT, HIT involved significantly higher mean and max HR during all bouts and training blocks. For both HIT and MAT, HR was significantly higher on the treadmill vs overground and HR significantly increased across sessions when expressed relative to baseline HR_peak_.
- Blood lactate was recorded after the treadmill bout in the middle session of 527 (94%) of 563 attended training weeks (HIT, 246/268 [92%]; MAT, 281/295 [95%]). Compared with MAT, HIT elicited a significantly higher lactate response overall.
- Step counts were recorded during 1,464 (87%) of the 1,675 attended treatment sessions (HIT, 712/800 [89%]; MAT, 752/875 [86%]). Compared with MAT, HIT produced significantly lower step counts during all training blocks. For both HIT and MAT, step counts significantly progressed across sessions, but significantly less so for HIT versus MAT.
- RPE was recorded after 1,632 (97%) of the 1,675 attended treatment sessions (HIT, 761/800 [95%]; MAT, 871/875 [∼100%]). Compared with MAT, HIT elicited significantly higher RPE overall.

### Adverse events

There were no serious adverse events related to study procedures and no significant between-group differences in any adverse event categories (Table 3). Four participants experienced serious adverse events during study participation that were all determined to be unrelated to study procedures, including: a seizure leading to temporary hospitalization (n=1, MAT group), a fall with hip fracture (n=1, MAT group), an episode of delirium leading to temporary hospitalization (n=1, MAT group) and a recurrent stroke (n=1, HIT group).

**Table 3.** Adverse Events (AEs). *Values are N participants with AE (total N AEs)*.

| **Table 3. Adverse Events (AEs).** *Values are N participants with AE (total N AEs).* |  |  |  |
| --- | --- | --- | --- |
|  | **HIT (N=27)**<br>800 sessions | **MAT (N=28)**<br>875 sessions | **HIT/MAT odds ratio [95% CI]\*** |
| **Any post-randomization AE** | 25 (109) | 23 (84) | 3.2 [0.5, 20.5] |
| -Grade 1 (mild) | 22 (80) | 22 (62) | 1.2 [0.3, 5.2] |
| -Grade 2 (moderate) | 14 (28) | 9 (19) | 2.7 [0.8, 9.1] |
| -Grade 3 (severe) | 1 (1) | 3 (3) | 0.3 [0.0, 3.4] |
| -Grade 4-5 (life threat-death) | 0 (0) | 0 (0) | 1 [NA] |
| -Pain/Soreness | 12 (25) | 11 (32) | 1.3 [0.4, 4.0] |
| -Fatigue | 4 (4) | 5 (6) | 0.8 [0.2, 3.6] |
| -Lightheadedness | 6 (17) | 7 (15) | 0.9 [0.2, 3.3] |
| -Fall | 9 (36) | 11 (14) | 0.7 [0.2, 2.3] |
| **Definitely treatment-related** | 13 (39) | 10 (29) | 1.7 [0.6, 5.2] |
| -Grade 1 (mild) | 11 (26) | 9 (24) | 1.5 [0.5, 4.7] |
| -Grade 2 (moderate) | 7 (13) | 4 (5) | 2.1 [0.5, 8.4] |
| -Grade 3-5 (severe-death) | 0 (0) | 0 (0) | 1 [NA] |
| -Pain/Soreness | 8 (15) | 7 (18) | 1.3 [0.4, 4.4] |
| -Fatigue | 2 (2) | 2 (2) | 1.1 [0.1, 8.7] |
| -Lightheadedness | 3 (10) | 4 (8) | 0.8 [0.1, 4.0] |
| -Fall | 3 (4) | 0 (0) | 8.2 [0.4, 167]<sup>†</sup> |
| **Possibly/not treatment-related** | 22 (70) | 19 (55) | 2.3 [0.6, 8.5] |
| -Grade 1 (mild) | 19 (54) | 17 (38) | 1.6 [0.5, 5.2] |
| -Grade 2 (moderate) | 10 (15) | 7 (14) | 2.4 [0.6, 9.7] |
| -Grade 3 (severe) | 1 (1) | 3 (3) | 0.3 [0.0, 3.4] |
| -Grade 4-5 (life threat-death) | 0 (0) | 0 (0) | 1 [NA] |
| -Pain/Soreness | 7 (10) | 8 (14) | 0.9 [0.3, 3.2] |
| -Fatigue | 2 (2) | 3 (4) | 0.7 [0.1, 5.9] |
| -Lightheadedness | 3 (7) | 5 (7) | 0.6 [0.1, 2.9] |
| -Fall | 9 (32) | 11 (14) | 0.7 [0.2, 2.3] |
| *Relative odds of a participant having an AE, adjusted for site and baseline gait dysfunction. <sup>†</sup>When AEs were only present in one group, continuity correction added 0.5 AEs to each group to permit (unadjusted) ratio calculation. |  |  |  |

### Primary outcome (6-minute walk test) changes

Within the HIT group, 6MWT distance significantly increased between each testing time point (Figure 5). Within the MAT group, the 6MWT did not significantly increase between any two consecutive time points but was significantly higher than PRE at the 8WK and 12WK time points.

**Figure 5.**
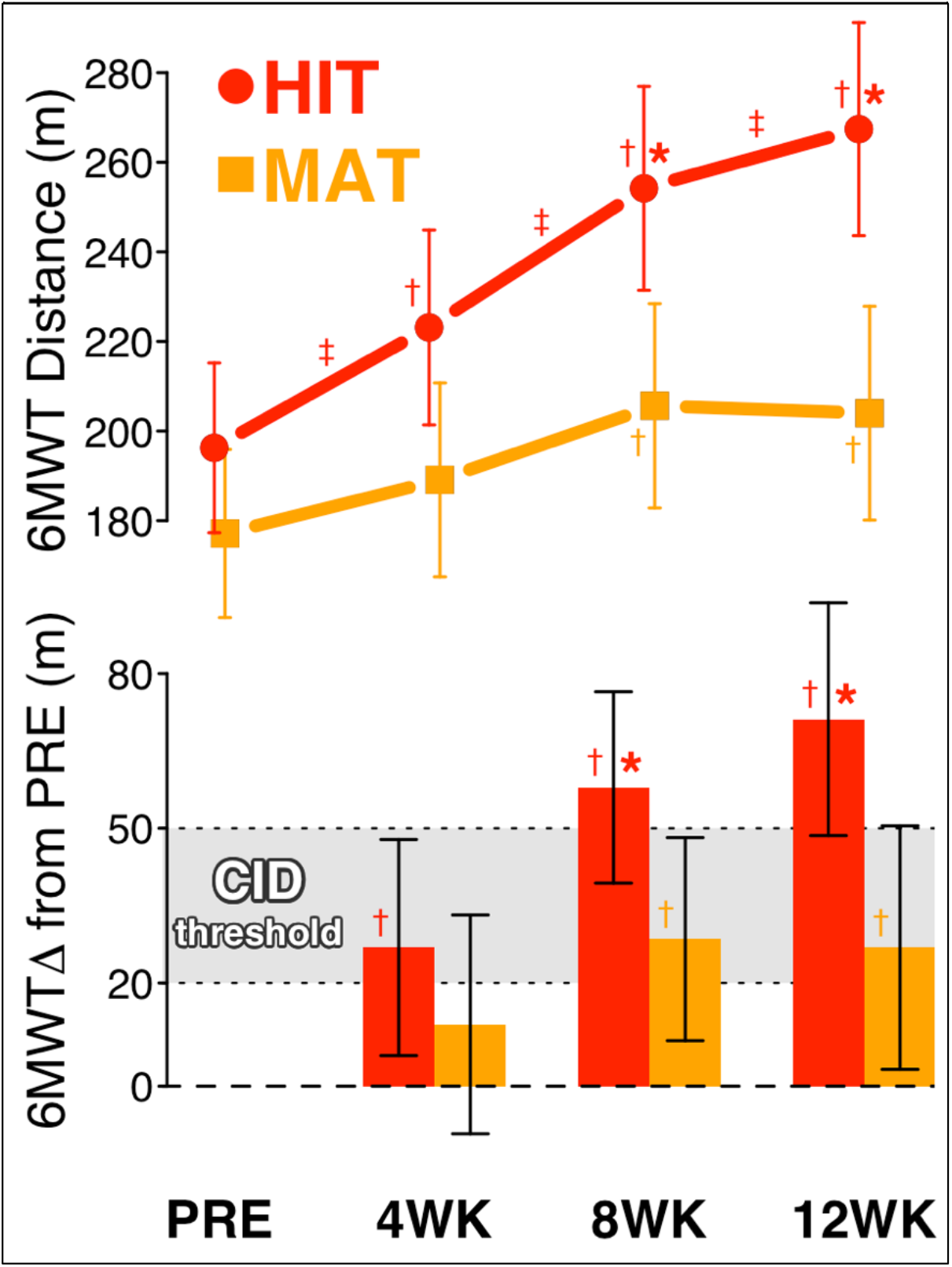
Primary outcome (6MWT) changes. Values are model estimates and error bars are 95% CI. ^‡^p<0.05 change (Δ) between time points ^†^p<0.05 Δ from baseline (PRE) *pFDR<0.05 between groups for Δ from PRE FDR, false discovery rate corrected across time points; 6MWT, 6-minute walk test; CID, clinically important difference (20-50m); WK, week

At the 4WK time point, there was no significant difference in 6MWT gains between groups (Table 4). At the 8WK and 12WK time points, the HIT group had improved significantly more than the MAT group by 29 m [95% CI: 5, 54 m] and 44 [14, 74] m, respectively.

**Table 4.** Primary and secondary outcome changes Baseline data presented as (covariate-adjusted) mean (SD) Change and difference data presented as mean difference [95% CI] *p<0.05 after false discovery rate-correction across time points & measures (primary & secondary) PROMIS, Patient Reported Outcomes Measurement Information System

|  | HIT<br>(N=27) | MAT<br>(N=28) | HIT - MAT | p-value |
| --- | --- | --- | --- | --- |
| **Walking Capacity: 6-Minute Walk Distance, m (primary outcome)** |  |  |  |  |
| Baseline (PRE) | 196 (98) | 177 (99) | 19 [-31, 69] | 0.4438 |
| 4-Week Change | 27 [6, 48] | 12 [-9, 33] | 15 [-13, 42] | 0.2824 |
| 8-Week Change | 58 [39, 76] | 29 [9, 48] | 29 [5, 54] | 0.0215* |
| 12-Week Change | 71 [49, 94] | 27 [3, 50] | 44 [14, 74] | 0.0048* |

| Self-Selected Gait Speed: 10-meter Walk Test, m/s |  |  |  |  |
| --- | --- | --- | --- | --- |
| Baseline | 0.52 (0.20) | 0.49 (0.21) | 0.03 [-0.07, 0.13] | 0.5607 |
| 4-Week Change | 0.11 [0.06, 0.15] | 0.02 [-0.02, 0.07] | 0.08 [0.02, 0.15] | 0.0091* |
| 8-Week Change | 0.14 [0.08, 0.20] | 0.06 [0.00, 0.12] | 0.08 [0.00, 0.15] | 0.0426 |
| 12-Week Change | 0.19 [0.13, 0.25] | 0.06 [0.00, 0.12] | 0.13 [0.05, 0.20] | 0.0026* |
| Fastest Gait Speed: 10-meter Walk Test, m/s |  |  |  |  |
| Baseline | 0.70 (0.32) | 0.62 (0.32) | 0.08 [-0.08, 0.24] | 0.3257 |
| 4-Week Change | 0.22 [0.16, 0.28] | 0.01 [-0.05, 0.07] | 0.21 [0.13, 0.29] | <0.0001* |
| 8-Week Change | 0.24 [0.17, 0.32] | 0.09 [0.01, 0.17] | 0.15 [0.05, 0.25] | 0.0029* |
| 12-Week Change | 0.28 [0.19, 0.37] | 0.09 [-0.01, 0.18] | 0.20 [0.08, 0.32] | 0.0016* |
| Fatigue: PROMIS-Fatigue Scale, T score (lower score = less fatigue) |  |  |  |  |
| Baseline | 52.4 (8.6) | 50.8 (8.7) | 1.7 [-2.7, 6.0] | 0.4473 |
| 4-Week Change | -1.7 [-4.0, 0.6] | 0.0 [-2.3, 2.3] | -1.7 [-4.8, 1.3] | 0.2590 |
| 8-Week Change | -3.0 [-5.5, -0.5] | 1.0 [-1.6, 3.6] | -4.0 [-7.3, -0.7] | 0.0185* |
| 12-Week Change | -1.1 [-3.7, 1.5] | -0.1 [-2.7, 2.5] | -1.0 [-4.4, 2.5] | 0.5678 |
| Ventilatory Threshold Oxygen Consumption Rate (VO<sub>2</sub>), mL/kg/min |  |  |  |  |
| Baseline | 11.5 (4.0) | 10.9 (4.0) | 0.6 [-1.4, 2.6] | 0.5522 |
| 4-Week Change | 1.3 [0.4, 2.2] | 0.4 [-0.5, 1.3] | 0.9 [-0.4, 2.1] | 0.1609 |
| 8-Week Change | 2.0 [0.7, 3.3] | 1.7 [0.3, 3.1] | 0.3 [-1.4, 2.1] | 0.7209 |
| 12-Week Change | 1.9 [0.4, 3.4] | 1.4 [-0.2, 3.0] | 0.5 [-1.5, 2.6] | 0.6018 |
| Baseline data presented as (covariate-adjusted) mean (SD)<br>Change and difference data presented as mean difference [95% CI]<br>*p<0.05 after false discovery rate-correction across time points & measures (primary & secondary)<br>PROMIS, Patient Reported Outcomes Measurement Information System |  |  |  |  |

### Secondary outcome changes

Both groups exhibited significant increases in self-selected gait speed, fastest gait speed and ventilatory threshold VO_2_ at various time points relative to PRE (Table 4), with the HIT group showing significantly greater increases than MAT in gait speeds. Only HIT significantly decreased PROMIS-Fatigue scores and only at the 8WK time point, which was significantly different between groups.

### Other exercise test measures to facilitate interpretation and exploratory outcomes

Other exercise capacity outcomes showed significant increases in both groups at various time points (Table 5), with some measures increasing significantly more for HIT versus MAT at 4WK (GXT duration, peak GXT speed and peak GXT HR) or 12WK (peak GXT HR and HR_peak_). The verification test increased HR_peak_ by 2.7 [1.0, 4.4] bpm at baseline and 2.0 [1.3, 2.7] bpm across all time points. Baseline HR_peak_ was 87.0 ± 17.2% age-predicted maximal HR (unadjusted mean ± SD). Peak respiratory exchange ratio and rating of perceived exertion during the baseline GXT were 1.01 ± 0.11 and 15.2 ± 2.6, respectively, and did not significantly change over time in either group. Meanwhile, there were significant improvements in both groups for ventilatory threshold treadmill speed, metabolic cost of gait in the last 3 minutes of the GXT, and functional ambulation category score, with no significant differences between groups. Only the HIT group reported significant improvement in overall quality of life (lower EuroQoL misery score) and performance of usual activities (lower EuroQoL score for that item), and this was only at 4WK, with no significant between-group differences after FDR correction.

**Table 5.** Other exercise test measures to facilitate interpretation and exploratory outcomes

|  |  |  |  |  |
| --- | --- | --- | --- | --- |
|  | HIT<br>(N=27) | MAT<br>(N=28) | HIT - MAT | p-value |
| Graded Exercise Test (GXT) Duration, minutes |  |  |  |  |
| Baseline (PRE) | 9.6 (4.2) | 8.9 (4.2) | 0.7 [-1.4, 2.8] | 0.5212 |
| 4-Week Change | 1.8 [1.1, 2.4] | 0.6 [-0.1, 1.2] | 1.2 [0.3, 2.1] | 0.0078* |
| 8-Week Change | 2.5 [1.5, 3.4] | 1.6 [0.6, 2.5] | 0.9 [-0.3, 2.1] | 0.1537 |
| 12-Week Change | 3.3 [2.2, 4.4] | 2.2 [1.1, 3.4] | 1.1 [-0.4, 2.6] | 0.1350 |
| Peak Treadmill Speed During GXT, m/s |  |  |  |  |
| Baseline | 0.69 (0.36) | 0.64 (0.37) | 0.05 [-0.14, 0.23] | 0.6081 |
| 4-Week Change | 0.15 [0.09, 0.22] | 0.04 [-0.03, 0.11] | 0.11 [0.03, 0.20] | 0.0093* |
| 8-Week Change | 0.17 [0.09, 0.26] | 0.13 [0.04, 0.22] | 0.04 [-0.07, 0.16] | 0.4391 |
| 12-Week Change | 0.24 [0.14, 0.33] | 0.20 [0.10, 0.30] | 0.04 [-0.09, 0.16] | 0.5570 |
| Peak Oxygen Consumption Rate During GXT (VO<sub>2-peak</sub>), mL/kg/min |  |  |  |  |
| Baseline | 13.4 (4.4) | 13.0 (4.4) | 0.4 [-1.8, 2.6] | 0.7160 |
| 4-Week Change | 1.5 [0.5, 2.5] | 0.2 [-0.8, 1.2] | 1.3 [0.0, 2.6] | 0.0478 |
| 8-Week Change | 2.1 [0.8, 3.5] | 1.5 [0.0, 2.9] | 0.7 [-1.2, 2.5] | 0.4778 |
| 12-Week Change | 2.5 [0.8, 4.2] | 1.5 [-0.3, 3.3] | 1.1 [-1.2, 3.4] | 0.3562 |
| Peak Heart Rate During GXT, bpm |  |  |  |  |
| Baseline | 123.4 (21.1) | 128.2 (21.3) | -4.9 [-15.5, 5.8] | 0.3663 |
| 4-Week Change | 5.4 [0.9, 9.9] | -3.7 [-8.2, 0.7] | 9.1 [3.3, 15.0] | 0.0030* |
| 8-Week Change | 4.2 [-1.5, 9.9] | -2.2 [-8.2, 3.9] | 6.4 [-1.3, 14.0] | 0.0994 |
| 12-Week Change | 12.6 [6.5, 18.6] | -1.0 [-7.2, 5.2] | 13.6 [5.6, 21.6] | 0.0013* |
| Highest Heart Rate During GXT or Verification Test (HR<sub>peak</sub>), bpm |  |  |  |  |
| Baseline | 125.0 (22.1) | 131.4 (22.3) | -6.4 [-17.6, 4.8] | 0.2562 |
| 4-Week Change | 5.7 [0.2, 11.1] | -3.7 [-9.2, 1.7] | 9.4 [2.2, 16.5] | 0.0111 |
| 8-Week Change | 4.1 [-1.7, 10.0] | -3.4 [-9.6, 2.8] | 7.5 [-0.3, 15.4] | 0.0605 |
| 12-Week Change | 11.2 [4.7, 17.8] | -2.6 [-9.3, 4.0] | 13.9 [5.2, 22.5] | 0.0023* |

| **HR<sub>peak</sub>, % Age-Predicted Maximal HR** |  |  |  |  |
| --- | --- | --- | --- | --- |
| Baseline | 83.3 (18.4) | 90.0 (18.5) | -6.7 [-16.0, 2.6] | 0.1528 |
| 4-Week Change | 3.8 [0.2, 7.4] | -2.4 [-6.0, 1.2] | 6.2 [1.4, 11.0] | 0.0119 |
| 8-Week Change | 3.4 [-0.7, 7.4] | -2.1 [-6.4, 2.2] | 5.4 [0.0, 10.9] | 0.0497 |
| 12-Week Change | 7.8 [3.1, 12.6] | -2.2 [-7.0, 2.7] | 10.0 [3.7, 16.3] | 0.0026* |
| **Peak Respiratory Exchange Ratio During GXT (VCO<sub>2</sub>/VO<sub>2</sub>)** |  |  |  |  |
| Baseline | 1.01 (0.12) | 1.02 (0.12) | -0.01 [-0.07, 0.05] | 0.7949 |
| 4-Week Change | -0.01 [-0.04, 0.02] | -0.03 [-0.06, 0.00] | 0.01 [-0.03, 0.05] | 0.5181 |
| 8-Week Change | 0.03 [-0.02, 0.07] | -0.02 [-0.06, 0.03] | 0.04 [-0.01, 0.10] | 0.1432 |
| 12-Week Change | 0.00 [-0.04, 0.04] | -0.01 [-0.05, 0.03] | 0.00 [-0.05, 0.06] | 0.8689 |
| **Rating of Perceived Exertion (RPE) from GXT, 6-20** |  |  |  |  |
| Baseline | 14.2 (2.5) | 15.1 (2.5) | -0.9 [-2.1, 0.4] | 0.1655 |
| 4-Week Change | 0.4 [-1.0, 1.8] | -0.5 [-1.9, 0.8] | 1.0 [-0.8, 2.7] | 0.2796 |
| 8-Week Change | 0.7 [-0.5, 1.9] | 0.1 [-1.1, 1.3] | 0.6 [-1.0, 2.1] | 0.4559 |
| 12-Week Change | 0.9 [-0.2, 2.0] | 0.6 [-0.5, 1.6] | 0.3 [-1.0, 1.7] | 0.6185 |
| **Ventilatory Threshold Treadmill Speed, m/s** |  |  |  |  |
| Baseline | 0.60 (0.32) | 0.53 (0.33) | 0.07 [-0.09, 0.23] | 0.3972 |
| 4-Week Change | 0.13 [0.07, 0.18] | 0.06 [0.00, 0.11] | 0.07 [-0.01, 0.14] | 0.0691 |
| 8-Week Change | 0.16 [0.09, 0.23] | 0.12 [0.05, 0.19] | 0.04 [-0.05, 0.13] | 0.3615 |
| 12-Week Change | 0.21 [0.13, 0.28] | 0.19 [0.11, 0.27] | 0.02 [-0.08, 0.12] | 0.7148 |
| **Metabolic Cost of Treadmill Gait During Last 3 Minutes of GXT, mL O<sub>2</sub>/kg/m** |  |  |  |  |
| Baseline | 0.44 (0.19) | 0.47 (0.19) | -0.03 [-0.12, 0.07] | 0.5372 |
| 4-Week Change | -0.08 [-0.13, -0.03] | -0.05 [-0.10, 0.00] | -0.03 [-0.10, 0.03] | 0.3293 |
| 8-Week Change | -0.10 [-0.14, -0.05] | -0.08 [-0.13, -0.03] | -0.01 [-0.08, 0.05] | 0.6751 |
| 12-Week Change | -0.12 [-0.17, -0.06] | -0.11 [-0.17, -0.06] | 0.00 [-0.07, 0.06] | 0.9025 |
| **Metabolic Cost of Treadmill Gait During Last 3 Minutes of Shortest GXT, mL O<sub>2</sub>/kg/m** |  |  |  |  |
| Baseline | 0.45 (0.18) | 0.48 (0.18) | -0.03 [-0.12, 0.06] | 0.5095 |
| 4-Week Change | -0.01 [-0.04, 0.02] | -0.01 [-0.04, 0.02] | 0.00 [-0.04, 0.04] | 0.9400 |
| 8-Week Change | -0.02 [-0.06, 0.02] | -0.04 [-0.08, 0.00] | 0.02 [-0.03, 0.08] | 0.4346 |
| 12-Week Change | -0.01 [-0.06, 0.03] | -0.04 [-0.08, 0.01] | 0.02 [-0.04, 0.08] | 0.4298 |
| **Gait Stability: Functional Ambulation Category, 0-4** |  |  |  |  |
| Baseline | 3.16 (0.58) | 3.19 (0.58) | -0.03 [-0.32, 0.26] | 0.8305 |
| 4-Week Change | -0.02 [-0.24, 0.20] | 0.17 [-0.05, 0.39] | -0.19 [-0.48, 0.10] | 0.1950 |
| 8-Week Change | 0.22 [-0.03, 0.46] | 0.31 [0.06, 0.56] | -0.09 [-0.42, 0.24] | 0.5727 |
| 12-Week Change | 0.26 [0.02, 0.50] | 0.27 [0.03, 0.51] | -0.01 [-0.33, 0.31] | 0.9475 |
| **Activities-Specific Balance Confidence (ABC) Scale Score, 0-100** |  |  |  |  |
| Baseline | 59.6 (18.9) | 63.0 (19.1) | -3.4 [-12.9, 6.2] | 0.4815 |
| 4-Week Change | -2.7 [-7.2, 1.7] | -4.6 [-9.1, -0.1] | 1.9 [-4.0, 7.7] | 0.5266 |
| 8-Week Change | 1.6 [-3.1, 6.4] | -1.8 [-6.7, 3.1] | 3.5 [-2.8, 9.7] | 0.2757 |
| 12-Week Change | 2.5 [-2.7, 7.7] | 0.6 [-4.7, 5.9] | 1.9 [-5.0, 8.8] | 0.5758 |
| **EuroQOL-5D-5L Misery Score, 5-25 (higher score = worse self-reported health problems)** |  |  |  |  |
| Baseline | 11.0 (2.7) | 10.7 (2.7) | 0.3 [-1.1, 1.7] | 0.6669 |
| 4-Week Change | -0.9 [-1.8, -0.1] | -0.2 [-1.1, 0.7] | -0.7 [-1.9, 0.4] | 0.2158 |
| 8-Week Change | -0.6 [-1.5, 0.3] | -0.4 [-1.3, 0.5] | -0.2 [-1.3, 1.0] | 0.7859 |
| 12-Week Change | -0.1 [-1.0, 0.8] | -0.6 [-1.5, 0.3] | 0.5 [-0.7, 1.7] | 0.3777 |
| **EuroQOL-5D-5L Mobility Score, 1-5 (higher score = worse self-reported problems)** |  |  |  |  |
| Baseline | 2.56 (0.86) | 2.51 (0.87) | 0.05 [-0.38, 0.49] | 0.8081 |
| 4-Week Change | -0.17 [-0.58, 0.24] | -0.15 [-0.56, 0.26] | -0.01 [-0.55, 0.52] | 0.9573 |
| 8-Week Change | -0.17 [-0.60, 0.27] | -0.13 [-0.58, 0.32] | -0.04 [-0.62, 0.54] | 0.8921 |
| 12-Week Change | -0.23 [-0.64, 0.18] | -0.24 [-0.65, 0.17] | 0.01 [-0.53, 0.54] | 0.9824 |
| **EuroQOL-5D-5L Usual Activities Score, 1-5 (higher score = worse self-reported problems)** |  |  |  |  |
| Baseline | 2.53 (0.88) | 2.23 (0.89) | 0.30 [-0.15, 0.74] | 0.1822 |
| 4-Week Change | -0.41 [-0.71, -0.11] | 0.02 [-0.28, 0.31] | -0.43 [-0.82, -0.04] | 0.0320 |
| 8-Week Change | -0.25 [-0.63, 0.14] | 0.11 [-0.29, 0.52] | -0.36 [-0.88, 0.15] | 0.1615 |
| 12-Week Change | -0.16 [-0.57, 0.24] | -0.07 [-0.48, 0.34] | -0.09 [-0.63, 0.44] | 0.7262 |
| Baseline data presented as (covariate-adjusted) mean ± SD |  |  |  |  |
| Change and difference data presented as mean difference [95% CI] |  |  |  |  |
| *p<0.05 after false discovery rate-correction across timepoints & all measures |  |  |  |  |
| PROMIS, Patient Reported Outcomes Measurement Information System |  |  |  |  |

### Primary outcome changes stratified by baseline gait dysfunction subgroup

Among the 41 participants (20 HIT, 21 MAT) with mild/moderate baseline gait dysfunction (self-selected speed 0.4-1.0 m/s), 6MWT distance significantly increased in both the HIT and MAT groups, with significantly greater improvement in HIT at 12WK (Figure 6, Table 6). Among the 14 participants (7 HIT, 7 MAT) with severe baseline gait dysfunction (self-selected speed <0.4 m/s), 6MWT distance significantly increased in the HIT group only, but with no significant difference between groups.

**Table 6.** Primary outcome (6-Minute Walk Distance) changes stratified by baseline gait dysfunction

| **Mild/Moderate Baseline Dysfunction** (gait speed 0.4-1.0 m/s; N=41) |  |  |  |  |
| --- | --- | --- | --- | --- |
|  | **HIT**<br>**(N=20)** | **MAT**<br>**(N=21)** | **HIT - MAT** | **p-value** |
| Baseline (PRE) | 305 (93) | 285 (93) | 21 [-38, 79] | 0.4816 |
| 4-Week Change | 31 [8, 55] | 20 [-2, 42] | 11 [-21, 43] | 0.4960 |
| 8-Week Change | 72 [50, 93] | 43 [24, 62] | 29 [0, 57] | 0.0490 |
| 12-Week Change | 88 [62, 114] | 44 [20, 67] | 44 [9, 79] | 0.0142* |
| **Severe Baseline Dysfunction** (gait speed <0.4 m/s; N=14) |  |  |  |  |
|  | **HIT**<br>**(N=7)** | **MAT**<br>**(N=7)** | **HIT - MAT** | **p-value** |
| Baseline (PRE) | 86 (93) | 71 (93) | 15 [-85, 115] | 0.7656 |
| 4-Week Change | 26 [-12, 64] | -1 [-42, 40] | 27 [-29, 83] | 0.3348 |
| 8-Week Change | 45 [12, 77] | 14 [-29, 56] | 31 [-22, 85] | 0.2464 |
| 12-Week Change | 55 [15, 95] | 10 [-40, 60] | 45 [-19, 109] | 0.1656 |

Baseline data presented as (covariate-adjusted) mean ± SD Change and difference data presented as mean difference [95% CI] \*p<0.05 after false discovery rate-correction across time points

**Figure 6.**
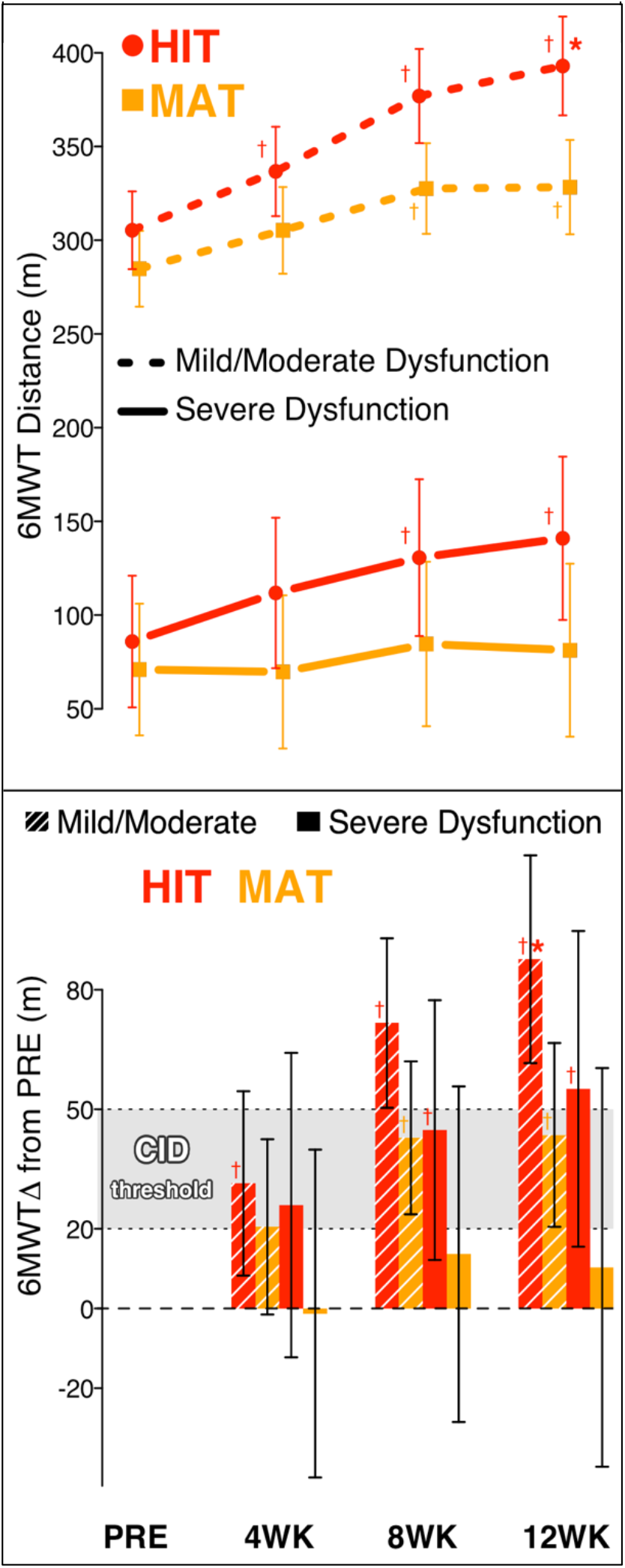
Primary analysis stratified by baseline gait dysfunction. Values are model estimates and error bars are 95% CI. Mild/moderate dysfunction, gait speed 0.4-1.0 m/s; severe dysfunction, <0.4 m/s. _†_p<0.05 change (Δ) from baseline (PRE) *pFDR<0.05 between groups for Δ from PRE FDR, false discovery rate corrected across time points; 6MWT, 6-minute walk test; CID, clinically important difference; WK, week

### Sensitivity of primary results to missing data assumptions

When assuming that participants who dropped out due to an adverse event had no mean improvement (rather than the common “missing at random” assumption), both groups still showed significant improvements in 6MWT distance and the direction of the between-group differences remained unchanged, but the between-group effect sizes were smaller and no longer statistically significant (Table 7).

**Table 7.**
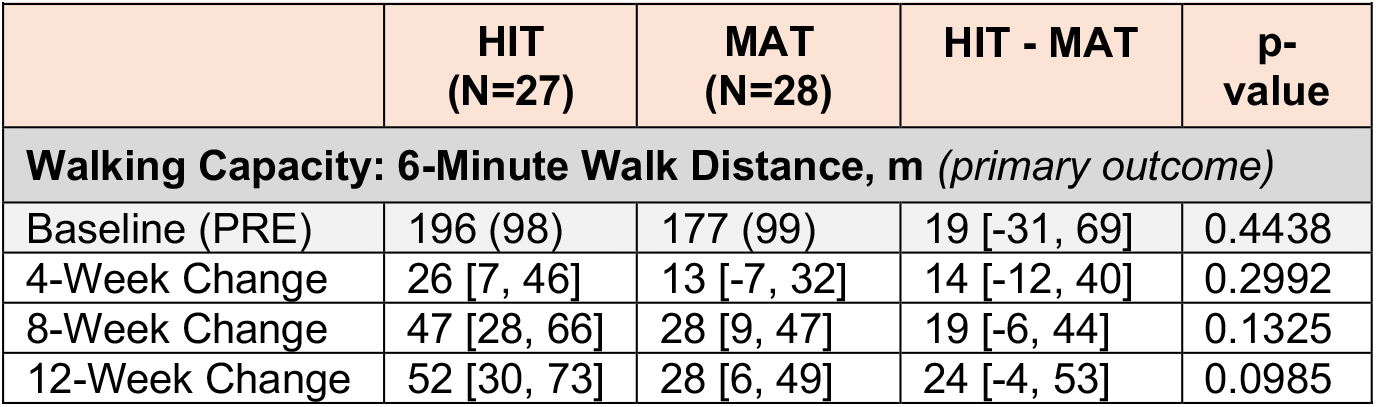
Estimated 6-minute walk test changes when assuming no mean improvement in cases of adverse-event-related dropout

|  | **HIT**<br>**(N=27)** | **MAT**<br>**(N=28)** | **HIT - MAT** | **p-value** |
| --- | --- | --- | --- | --- |
| **Walking Capacity: 6-Minute Walk Distance, m (primary outcome)** |  |  |  |  |
| Baseline (PRE) | 196 (98) | 177 (99) | 19 [-31, 69] | 0.4438 |
| 4-Week Change | 26 [7, 46] | 13 [-7, 32] | 14 [-12, 40] | 0.2992 |
| 8-Week Change | 47 [28, 66] | 28 [9, 47] | 19 [-6, 44] | 0.1325 |
| 12-Week Change | 52 [30, 73] | 28 [6, 49] | 24 [-4, 53] | 0.0985 |

## DISCUSSION

Results of this multi-center trial gave proof of concept that the vigorous training intensity of HIT is a crucial dosing parameter for locomotor exercise in chronic stroke. Findings also indicated that locomotor HIT can produce significant and meaningful gains in walking capacity in 4 weeks, but that a training duration of at least 12 weeks is needed to maximize immediate gains in walking capacity with this intervention. Our primary hypothesis that HIT would improve walking capacity significantly more than MAT after 4 weeks of training was not supported (mean 6MWT change in HIT vs MAT: +15 meters [95% CI: -13-42]). However, the gap between groups continued to widen over time, and HIT elicited significantly greater improvement in walking capacity than MAT after 8 weeks (+29 [5-54] meters) and 12 weeks of training (+44 [14-74] meters) in the prespecified primary analysis, supporting the secondary hypotheses. Among the secondary and exploratory outcomes, HIT also elicited significantly greater improvements than MAT in gait speed (self-selected and fastest), fatigue, and exercise capacity (GXT duration, peak GXT speed and peak HR) at various time points after adjustment for multiple comparisons (Tables 4 & 5). In addition, intensive safety monitoring was successful at identifying expected AEs and results suggest that post-stroke HIT continues to be sufficiently safe for further study.

Within the MAT group, the mean 6MWT gain of +27 meters [95% CI: 3-50] after 12 weeks of training was comparable to many similar previous studies in chronic stroke,^25,26,29-31,33,115^ and exceeded the minimally important difference of +20 meters.^86^ This confirms that MAT was successfully implemented in this study and has some value for improving walking capacity in this population. Surprisingly, our results suggest that most outcomes reached an apparent plateau after just 8 weeks of MAT, which may not support the longer 3-6 month training durations used in most previous stroke trials focused on MAT. However, there are prior indications that outcomes may continue to slowly improve between 3-6 months of MAT.^28,33^ In addition, it is plausible that longer training durations could result in more sustained gains in walking capacity or greater long-term improvements in cardiovascular-metabolic health, but this has not been thoroughly tested.

Within the HIT group, the mean 6MWT gain after 4 weeks of training was comparable to that found in prior single-site 4-week pilot testing,^63^ confirming successful multi-center implementation. As hypothesized, gains in walking capacity and other outcomes were even larger after 8 and 12 weeks of HIT, with no apparent plateau. These findings suggest that most prior post-stroke HIT studies have used insufficient training durations, and that HIT durations of at least 12 weeks should be considered in the future. Outpatient therapy durations longer than 12 weeks (36 sessions) are not likely to be routinely feasible in the current model for clinical rehabilitation due to reimbursement constraints^116-118^ and adherence issues.^119,120^ Thus, a 12-week duration for clinician-led HIT program may be an optimal standard to target.

Our prespecified subgroup analysis suggested that the vigorous training intensity of HIT may have been particularly important for participants with severe baseline gait dysfunction (self-selected gait speed <0.4 m/s^62,63^). For these individuals, MAT elicited mean 6MWT gains less than the minimally important difference of +20 meters^86^ (+10 [-40-60] meters). Previous research also found minimal response to MAT among stroke survivors with severe gait dysfunction,^43,62^ leading to prior suggestions that these patients may not be good candidates for gait training.^62^ However, in the current study, participants with severe gait dysfunction increased mean 6MWT distance by +55 [15-95] meters with HIT, which is above the +50-meter threshold for a large clinically-meaningful gain.^86^ This study was not powered to detect significant differences between HIT and MAT within the severe dysfunction subgroup (N=14, p=0.17) or to assess within-group changes with sufficient precision to rule meaningful benefits in or out with confidence, so these findings should be taken as preliminary. However, HIT did show significantly greater improvement than MAT in the larger mild/moderate dysfunction subgroup (+88 [62-114] vs +44 [20-67], N=41, p=0.01), suggesting that vigorous training intensity could be optimal regardless of baseline dysfunction.

The intensity-dependent improvement (decrease) in PROMIS-Fatigue T scores after 8 weeks of training (HIT -3.0 [-5.5, -0.5], MAT 1.0 [-1.6, 3.6], p=0.02) is especially noteworthy because identifying a treatment for post-stroke fatigue has been highlighted as a research priority.^121^ This finding is consistent with prior neurophysiologic evidence showing that post-stroke fatigue is related to lower corticomotor excitability^122^ and that HIT elicits significantly greater acute increases in corticomotor excitability than MAT.^123^ Interestingly, participants randomized to HIT reported this significant decrease in fatigue during a time period when they were coming to the research sites multiple times per week for training sessions that commonly *induced* temporary fatigue. One possible explanation for this apparent paradox is that the acute central motor activation effects of post-stroke HIT appear to counterbalance or exceed the effects of fatigue.^123^ It is also possible that the greater longitudinal increases in walking capacity and exercise tolerance with HIT may have reduced fatigue with everyday activities, especially when coupled with significant reductions in the metabolic cost of gait. Oddly, HIT group improvements and between-group differences in fatigue were only statistically significant after 8 weeks of training and not after 4 or 12 weeks of training. Yet all these effect estimates were in the same direction, suggesting that the lack of significance at the 4 and 12-week timepoints may have been due to insufficient statistical power, rather than an exclusive fatigue benefit at only the 8-week time point. Conversely, it is always possible that the significant improvement in fatigue observed after 8 weeks of HIT was due to chance, but such improvement was also observed in a prior 4-week pilot study,^63^ suggesting otherwise. Confirming these findings in a larger trial with follow up testing is warranted.

Both groups showed significant improvement in aerobic capacity (ventilatory threshold VO_2_) and VO_2_-peak. The lack of significant between-group differences in aerobic capacity gains suggests that the greater improvement in walking capacity with HIT vs MAT may have been primarily driven by neuromotor rather than cardiopulmonary adaptations. Although, it is also possible that this lack of significant between-group differences was due to greater measurement error for VO_2_-based outcomes (ventilatory threshold and peak) or confounding effects of gait efficiency changes on VO_2_ during exercise testing (improved gait efficiency decreases measured VO_2_).^89,90,93,94^ As expected, the symptom-limited GXT was primarily terminated by non-cardiorespiratory factors on average (respiratory exchange ratio < 1.05, RPE < 17, HR ≲ 90% age-predicted max), so GXT duration and peak GXT measures cannot be interpreted as specific measures of cardiopulmonary (aerobic) capacity, but can still be interpreted as measures of *exercise capacity*, which can be influenced by neuromotor function, cardiopulmonary capacity and other factors (e.g. motivation).

Importantly, significant improvements in the metabolic cost of gait and gait stability for both groups suggest that compensatory movements and compromised stability were not the primary mechanisms by which individuals achieved gains in walking capacity with training. However, it did appear that gains in efficiency were primarily related to gains in gait speed, since efficiency at matched speeds did not significantly change.

## Strengths and limitations

Most recent post-stroke HIT studies have focused on improving aerobic fitness among persons with mild stroke or transient ischemic attack who had unimpaired, near-normal or unreported baseline walking function.^41,42,44,46,47,124,125^ Thus, one important strength of this trial is its relevance to clinical rehabilitation, since we focused on improving walking capacity as the primary outcome and recruited a sample with impaired gait (self-selected speed, 0.63 ± 0.31 m/s, which was 49 ± 24% of normative predicted values) for therapist-led training. Another major strength of this trial is our systematic AE data collection with blinded adjudication, which is rare in the rehabilitation literature, but critical for understanding the risk/benefit ratio of an intervention.

In terms of rigor, this study used randomization, concealed allocation, assessor blinding and intent-to-treat analysis to minimize risk of bias^113^ and only had 10% missing outcome data. We also added the rigor of multi-center implementation earlier than usual in the rehabilitation research continuum,^126^ increasing confidence in reproducibility and generalizability of the results. In addition, we included more rigorous assessment than many clinical trials about how missing data might influence the results.^113^ Further, we successfully implemented several novel technological methods to enhance protocol fidelity, including automated real-time prompts, calculations and feedback from the direct electronic data entry system during all visits, plus automated bout timing, recording & intensity guidance from a fully wireless HR monitoring system during treatment visits.

The primary limitation to this trial was its relatively modest sample size (N=55), since it was designed to be an early proof of concept trial with a secondary aim of optimizing training duration dosing. This meant that we were likely underpowered to detect all but very large differences in AEs between HIT and MAT. The small sample size is also probably why the primary findings were not fully robust to different plausible assumptions about the true values of the missing data (Table 7). This means that the findings should not be viewed as definitive and that they require verification in a larger trial. This sensitivity analysis also validates the general intuition that larger samples are needed to have more confidence in the results of clinical trials.

Another important limitation was the lack of follow up testing to assess sustained effects after treatment ended. This also makes it more challenging to interpret the results of the questionnaires (PROMIS-Fatigue, ABC scale and EuroQOL-5D-5L) since they were asking participants to report about time periods in which they were engaged in the study treatment at 4WK, 8WK and 12WK, but asking about a time period when participants were largely just participating in normal daily activities at PRE. We attempted to mitigate this issue by specifically asking participants to answer questions based on their usual activities outside of the study but are skeptical that this strategy was completely successful. Thus, a larger trial with post-treatment follow-up testing is needed to confirm and expand on these promising results.

## Conclusions

For locomotor exercise among stroke survivors with chronic gait dysfunction, vigorous training intensity appears to be significantly better than moderate intensity for eliciting immediate improvements in walking capacity and other outcomes. With this vigorous intensity locomotor exercise, 12 weeks of training produces greater gains than shorter durations. Preliminary results among patients with severe baseline gait dysfunction (self-selected speed <0.4 m/s) suggest that a vigorous training intensity could possibly be necessary for this subgroup to have meaningful benefits.

## Data Availability

Data produced in the present study will be deposited into the National Institute of Child Health and Human Development (NICHD) Data and Specimen Hub (DASH) repository after this manuscript is accepted for publication.

## Trial Registration

ClinicalTrials.gov Identifier: NCT03760016. First posted: November 30, 2018. https://clinicaltrials.gov/ct2/show/NCT03760016

## Acknowledgements

The authors thank Linda D’Silva, Rachel Gleason, Bill Hendry, Chun Kai-Huang, Katie Libby, Babette Northrop, Jenna Octavio, Jaclyn Reddington, Caio Sarmento, Eric Vidoni, Kaylah Williamson and Tamara Wright for assistance with data collection and Kelly Laipply for cardiology consultation. We also thank T George Hornby, Michael W O’Dell and Bin Huang for serving on the data safety monitoring board.

